# Profiling the effect of micronutrient levels on vital cardiac markers

**DOI:** 10.1101/2023.04.19.23288794

**Authors:** Hari Krishnan Krishnamurthy, Swarnkumar Reddy, Vasanth Jayaraman, Karthik Krishna, Qi Song, Tianhao Wang, Kang Bei, John J. Rajasekaran

## Abstract

Cardiovascular diseases (CVD) are among the most preventable chronic disorders accounting for about one-third of general mortality around the globe. Micronutrients have been shown to have a significant impact on cardiovascular health. Micronutrients have been looked at as the most adoptable lifestyle choice which could reduce the burden of disease around the world. In this context, it is important to study the levels of micronutrients and see their correlation to cardiac disease biomarkers. The present study, has attempted to investigate the relationship between the diverse class of micronutrients and serum levels of the key lipids and lipoproteins. A retrospective analysis was carried out between the serum levels of micronutrients and vital cardiovascular markers. The study was carried out in a group of 358 individuals tested for the Cardio Health and Micronutrients Panel at Vibrant America Clinical Laboratory. The study population was categorized based on the serum concentration of lipids and lipoproteins into 3 groups ‘Low’ ‘Normal’ and ‘High’ and the levels of micronutrients were compared among these groups. The results revealed a significant association of several cardiovascular markers with vitamins including Vit D, Vit E, Vit K, and minerals including zinc, iron, calcium, magnesium, and amino acids including leucine, isoleucine, and valine. Quantitative analysis by Pearson’s correlation exhibited a negative correlation of asparagine with serum levels of cholesterol and LDL. Amino acids such as cysteine, isoleucine, and valine were found to have a significant negative correlation with HDL. A positive correlation was observed between valine and serum levels of LDL and Apo B. Vitamins such as Vit A, Vit D3, Vit E, and Vit K1 were found to have a strong positive correlation with levels of total cholesterol and triglycerides. The study summarizes micronutrients and modulation of several lipid markers which are critical for the management of cardiovascular diseases. Micronutrients such as vitamins B1, B3, asparagine, and glutamine have a strong positive association, and fat-soluble vitamins, and BCAA has a strong negative association with cardiovascular health.

## Introduction

Micronutrients are the most essential nutrients for a whole range of physiological functioning of a living system. Dietary deficiency of micronutrients leads to pathogenesis, progression, and morbidity in various critical clinical disorders including cardiovascular diseases (CVD) [1]. The pathobiology of CVD remains complex and it is generally regarded to be a result of various genetic predispositions interacting with environmental factors. More than 40% of the CVD are related to nutritional factors and more than 90% of such cases were attributed to preventable factors with nutrition as a major determinant [2]. Atherosclerosis and hypertension are the predominant chronic cardiovascular diseases that are profoundly preventable with a multi-component strategy including nutrient, diet, and lifestyle interventions [1]. Atherosclerosis, characterized by the chronic inflammation of arteries affecting the function of the heart results from the deposition of fats, cholesterol, and other substances on the inner lining of the artery eventually resulting in cardiovascular disease or heart failure [3]. It is estimated that ≥ 2% of the adult population has atherosclerosis which tends to increase up to 10% or higher in people above 70 years and continues to increase with the age of the population [4]. Hypertension is another predominant CVD, which is clinically referred to as a chronic sustained increase in arterial blood pressure. Hypertension is the most widespread CVD with more than 35% of the population affected globally, A report from the World health organization stated this might increase beyond 50% in 2025 [3]. Cardiac disease, being a complex multifunctional etiology, the precise mechanism of pathophysiology remains unclear in both atherosclerosis and hypertension. Atherosclerosis is characterized by the impaired flow of blood and hypertension is characterized by a chronic increase in blood pressure, both these conditions result in abnormal cardiac functioning and vascular damage [5].

Cardiovascular diseases include a wide spectrum of risk factors such as genetic, behavioral, and environmental factors. In most cases CVDs commonly result from the interplay of multiple risk factors, common classical risk factors include age, any pre-existing medical illness such as diabetes, hypertension, predisposition genetics, or behavioral factors like smoking. Among these risk factors, environmental factors are the most easily modifiable and play a pivotal role in cardiovascular health. The environmental factors are the least investigated risk factors and are noted as the most preventable risk factors of CVDs. Environmental factors include the intake of micronutrients such as vitamins, minerals, and amino acids. The micronutrients include organic compounds such as Vit A, Vit C, Vit D, Vit E, and Vit K, dietary minerals including calcium, sodium, potassium, magnesium, etc, and trace elements like iron, chromium, copper, selenium, and zinc [6, 7, 8, 9]. Several clinical reports have shown a significant correlation between the deficiency of one or more micronutrients and cardiovascular diseases. However, those reports were limited by the diversity of the micronutrients studied. For instance, Rai et al. [10] evaluated the effect of two micronutrients copper and zinc on lipid profiles in healthy adults, and another study by Islam et al. [6] reported that no significant impact was observed by consumption of vitamin D with calcium on serum lipids. The current study aimed to broadly investigate the correlation between cardiovascular markers and the levels of 32 micronutrients. We also attempted to hypothesize the pathophysiology of micronutrient deficiency through which the cardio biomarkers could be affected.

## Material and methods

### Study population

A total of 358 candidates with a mean age of 48±15 years who were tested for cardiovascular panel and micronutrient panel between December 2020 and July 2021 at Vibrant America Clinical Laboratory were included in the study. The study was categorized as a retrospective analysis of observed clinical data and hence was exempted from formal ethical review by Western IRB (work order #1-1098539-1) (Washington USA). The study was conducted in a free-living general population with no clinical indications of abnormalities. The subjects were classified based on the serum levels of lipids and lipoproteins as detailed in Table 1.

**Table 1:**
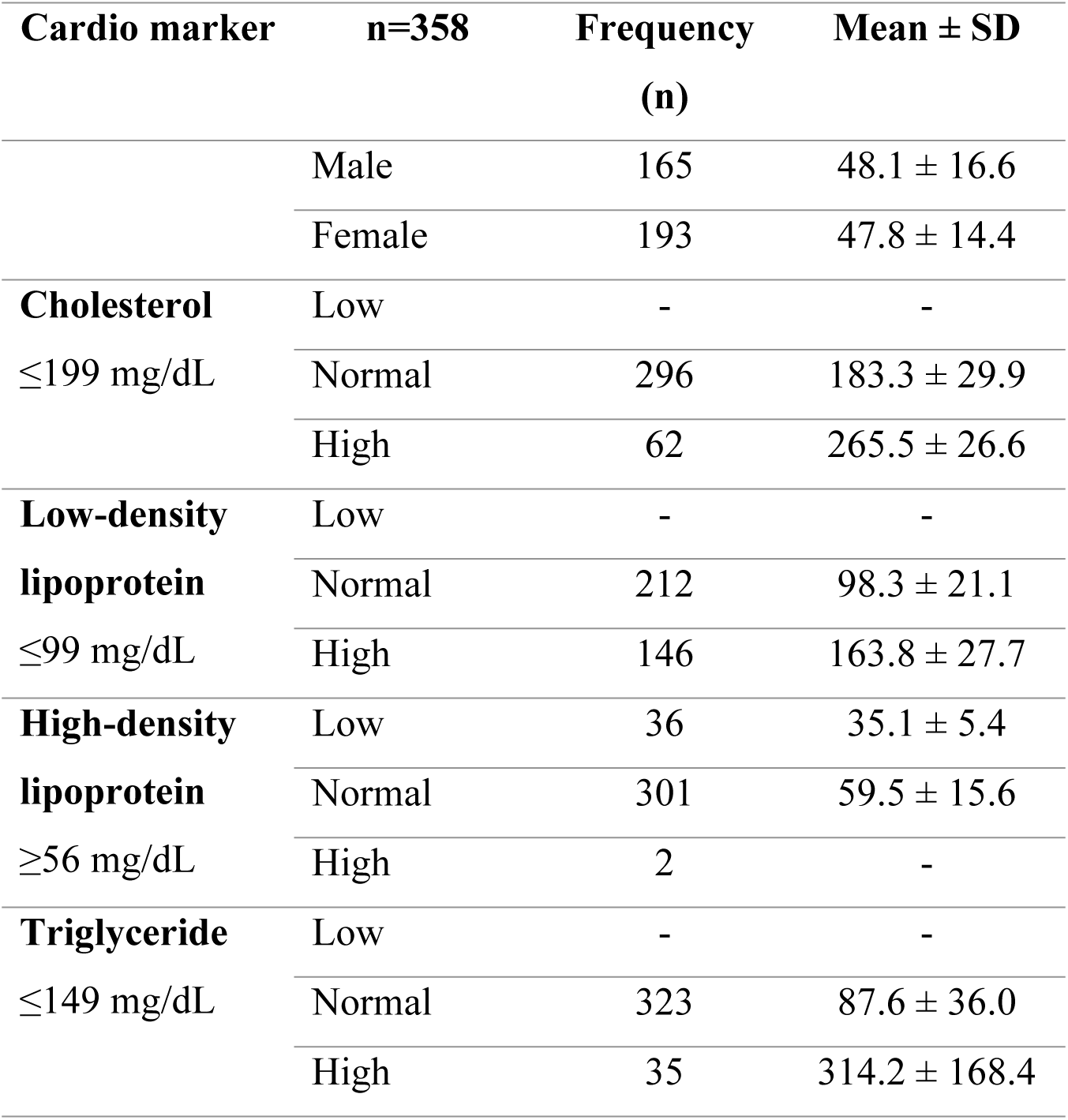
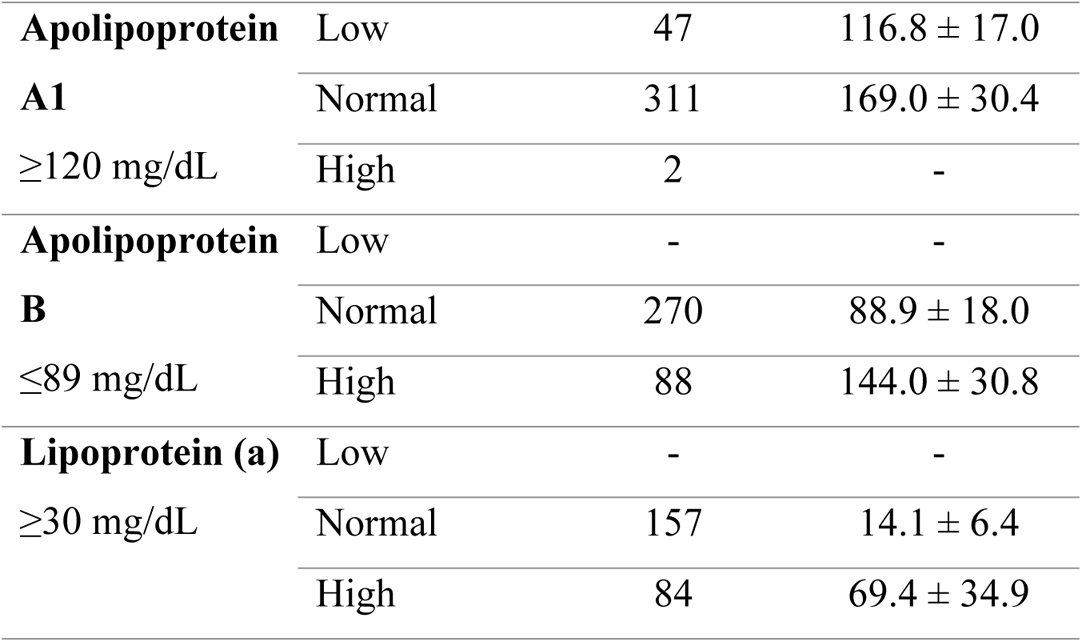
Serum Lipid, and Lipoprotein levels of candidates

### Cardiovascular Panel

Blood samples were processed for the separation of serum and further analyzed for a cardiovascular panel comprised of lipids (total cholesterol, LDL, HDL, and triglycerides), apolipoproteins (Apo A1, Apo B), and a lipoprotein marker lipoprotein (a). Total cholesterol was measured by the cholesterol dehydrogenase method via the Beckman Coulter AU680 analyzer. Serum levels of LDL, HDL, and triglycerides were measured by an enzymatic-colorimetric method using the Beckman Coulter AU680 analyzer. Other cardiovascular markers such as Apo A1, Apo B, and Lp (a) were also measured by a particle-enhanced immunoturbidimetric assay via Roche Cobas 6000 c 501 analyzers

### Micronutrient Panel

Serum levels of micronutrients were determined using Waters TQ-XS Tandem mass spectrometer coupled with LCMS, Waters GC-MS, and Perkin Elmer NexION ICP-MS using standard protocols.

### Statistical Analysis

Clinical data were subjected to retrospective analysis from de-identified subjects using Java for windows version 1.8.161. Non-parametric Mann-Whitney U test was used to compare the micronutrients with normal and altered lipid and lipoprotein concentrations. Pearson’s correlation was carried out to analyze the univariant relationship between serum lipids, lipoproteins, and micronutrients with significance set at *p*<0.05. All statistical analysis was performed using GraphPad Prism Version 7.00 and a descriptive statistic was used to define the continuous variables (mean ± SD, median, minimum and maximum).

## Results

The study group comprised 165 males and 193 females with a mean age group of 47.9 ± 15.4, table 1 shows the descriptive statistics of the lipid and lipoprotein profile of 358 subjects. The study aimed to evaluate the significant relationship of the wide range of micronutrients including vitamins, dietary minerals, trace elements, and organic compounds with cholesterol (total, LDL, HDL), triglycerides as well as apo A, apo B, and Lp (a) levels.

The subjects were classified based on the levels of serum lipids and lipoproteins. In the case of total cholesterol (TC), LDL, triglycerides, and Apo B the subjects were categorized based on lipid concentrations higher than the reference range and subjects within the reference range while in the case of HDL and apo A the subjects were divided into lipid concentrations lower than the reference range and subjects within the reference range. A significant statistical relationship was observed between various micronutrients and serum lipid and lipoprotein concentrations as shown in table 2. An increase in the serum levels of Vit E, Vit D3, and Mg were found to be statistically significant with increased levels of serum cholesterol and LDL. A decrease in serum levels of HDL was significantly affected by the serum levels of Vit D 25(OH), asparagine, glutamine, and serine. A high level of serum triglycerides is significantly associated with increased levels of various micronutrients including Vit A, Vit D 25(OH), Vit E, Vit K1, and amino acids such as glutamine, serine, isoleucine, valine, and leucine.

**Table 2:** Significant association of micronutrients with serum lipids and lipoprotein

| **CHOLESTEROL** |  |  |  |  |  |
| --- | --- | --- | --- | --- | --- |
|  | Greater than Reference |  | Within range (n=296) |  | P |
|  | Range (n=62) |  |  |  | (*p*<0.05) |
| Vitamin E | 16±5.7 | 14.4 (7.9-29.4) | 12.9±4.2 | 11.9 (5.3-30.7) | 0.0001 |
| Vitamin D3 | 1.1±0.2 | 1.1 (0.4-1.7) | 0.8±0.2 | 0.8 (0.3-1.6) | 0.0001 |
| Vitamin K1 | 2.1±1.5 | 1.5 (0.1-7.3) | 1.4±1.3 | 1.2 (0.05-13.9) | 0.0003 |
| Calcium | 9.8±0.3 | 9.8(9-10.6) | 9.6±0.4 | 9.6 (8.5-11.2) | 0.0001 |
| Magnesium | 2.2±0.1 | 2.2 (1.9-2.6) | 2.1±0.1 | 2.1 (1.0-2.6) | 0.0001 |
| **LOW-DENSITY LIPOPROTEIN** |  |  |  |  |  |
|  | Greater than Reference<br>Range (n=146) |  | Within range (n=212) |  | P<br>(*p*<0.05) |
| Vitamin E | 14.8±4.8 | 13.6(7.3-29.4) | 12.3±4.2 | 11.2(5.3-30.7) | 0.0001 |
| Vitamin D3 | 1.04±0.26 | 1.02(0.44-1.78) | 0.84±0.81 | 0.81(0.32-1.63) | 0.0001 |
| Iron | 108±33.3 | 108.1(35.5-201.8) | 99.1±41.4 | 91.4(25.3-238.9) | 0.0024 |
| Magnesium | 2.23±0.16 | 2.2(1.5-2.66) | 2.16±0.19 | 2.1(1.0-2.6) | 0.0002 |
| **HIGH-DENSITY LIPOPROTEIN** |  |  |  |  |  |
|  | Less than Reference<br>Range (n=36) |  | Within range (n=301) |  | P<br>(*p*<0.05) |
| Vitamin D 25 OH | 40.7±24.9 | 33.6(12.4-136) | 48.3±22.8 | 41.7(8.8-133) | 0.0091 |
| Asparagine | 105.6±42.6 | 110.1(34.1-220.6) | 53.9±11.6 | 52.9(28.1-108.0) | 0.0001 |
| Glutamine | 212.5±164.3 | 124.6(61.0-671.4) | 508.6±80.7 | 510.9(202.2-752.4) | 0.0001 |
| Serine | 128.0±36.2 | 124.7(61.0-282.9) | 149.4±34.2 | 145.6(59.6-274.5) | 0.0001 |
| **TRIGLYCERIDES** |  |  |  |  |  |
|  | Greater than Reference<br>Range (n=36) |  | Within range (n=323) |  | P<br>(*p*<0.05) |
| Vitamin E | 20.1±17 | 16.7(8.5-118.7) | 12.9±4.3 | 11.9(5.3-29.4) | 0.0001 |
| Vitamin A | 88.4±27.3 | 82(52.8-151.9) | 75.8±23.3 | 72.1(34.1-151.5) | 0.0061 |
| Vitamin K1 | 3.14±2.7 | 2.3(0.22-13.9) | 1.42±0.99 | 1.22(0.05-7.32) | 0.0001 |
| Vitamin D25 OH | 33.9±13.6 | 30.7(15.4-71.2) | 48.6±23.2 | 41.8(8.8-136) | 0.0001 |
| Glutamine | 463.7±107.6 | 472.9(118.7-608.3) | 508.4±78.7 | 507.6(202.2-752) | 0.0290 |
| Serine | 136.5±32.8 | 130.6(76.0-198.5) | 149.6±34.7 | 145.6(59.6-282.9) | 0.0368 |
| Carnitine | 32.6±16.5 | 31.5(11.6-118.7) | 26.5±7.2 | 26.6(4.1-50.5) | 0.0014 |
| Isoleucine | 84.0±27.5 | 84.2(35.7-142.7) | 67.1±19.1 | 64.6(24.0-139.7) | 0.0002 |
| Valine | 275.0±63.3 | 281.1(175.3-442.4) | 241.7±52.8 | 233.6(134.9-442.7) | 0.0020 |
| Leucine | 179.9±36.8 | 182.7(113.7-233.5) | 161.7±34.1 | 157.4(79.7-249.2) | 0.0033 |
| **APOLIPOPROTEIN A** |  |  |  |  |  |
|  | Less than Reference<br>Range (n=47) |  | Within range (n=323) |  | P<br>(*p*<0.05) |
| Vitamin E | 12.2±4.8 | 10.9(4.0-23.4) | 13.5±4.7 | 12.4(5.6-30.7) | 0.0369 |
| Vitamin D3 | 0.84±0.24 | 0.81(0.41-1.5) | 0.94±0.28 | 0.91(0.32-2.19) | 0.0198 |
| Asparagine | 51.0±10.6 | 49.4(34.1-79.1) | 54.5±12.1 | 53.3(28.1-113.0) | 0.0444 |
| Calcium | 9.4±0.3 | 9.5(8.7-10.4) | 9.6±0.39 | 9.6(8.5-11.1) | 0.0022 |
| Zinc | 0.69±0.19 | 0.64(0.45-1.50) | 0.71±0.15 | 0.69(0.39-2.23) | 0.0315 |
| Iron | 85.6±36.6 | 78.5(25.3-182.4) | 105.5±38.4 | 101.6(33.2-238.9) | 0.0015 |

**APOLIPOPROTEIN B**
|  | Greater than Reference<br>Range (n=87) |  | Within range (n=270) |  | P<br>(*p*<0.05) |
| --- | --- | --- | --- | --- | --- |
| Vitamin E | 16.0±5.5 | 15.3(7.3-29.0) | 12.5±4.1 | 11.7(4.0-30.7) | 0.0001 |
| Vitamin A | 82.4±23.7 | 77.7(34.1-145.8) | 75.8±24.2 | 72.1(37.1-151.9) | 0.0096 |
| Vitamin D3 | 0.87±0.24 | 0.83(0.32-1.6) | 1.09±0.30 | 1.08(0.44-2.19) | 0.0001 |
| Vitamin K1 | 1.5±1.3 | 1.2(0.05-13.9) | 1.8±1.4 | 1.4(0.10-7.3) | 0.0416 |
| Selenium | 146.4±24.0 | 144.8(93.4-210.8) | 142.3±30.3 | 139(95.7-360.4) | 0.0451 |
| Potassium | 4.5±0.35 | 4.5(3.6-5.6) | 4.5±0.40 | 4.49(3.1-6.22) | 0.0377 |
| Calcium | 9.7±0.3 | 9.7(8.7-10.6) | 9.6±0.3 | 9.6(8.5-11.1) | 0.0055 |
| Zinc | 0.73±0.11 | 0.72(0.39-1.0) | 0.70±0.17 | 0.68(0.44-2.23) | 0.0058 |
| Iron | 109.3±34.2 | 107.7(45.6-201.3) | 100.8±39.9 | 97.7(25.3-238.9) | 0.0239 |
| Magnesium | 2.2±0.1 | 2.2(1.9-2.9) | 2.1±0.19 | 2.1(1.0-2.6) | 0.0002 |
| Isoleucine | 72.7±19.9 | 70.7(34.0-121.4) | 67.6±20.7 | 64.2(24.0-142.7) | 0.0138 |
| Valine | 261.9±55.2 | 262.7(146.5-404.0) | 240.2±54.2 | 232.1(134.9) | 0.0004 |
| Leucine | 172±34.8 | 168.8(113.7-248.2) | 160.5±34.2 | 157.4(79.7-249.2) | 0.0138 |
| Serine | 140.6±33.0 | 135.4(75.0-209.7) | 151.1±34.7 | 147.2(59.6-282.9) | 0.0189 |

Decreased levels of serum Apo A are significantly associated with decreased serum levels of Vit E, Vit D3, asparagine, and minerals such as calcium, zinc, and iron. The elevated levels of Apo B associated with increasing the serum LDL and non-HDL fats are found to be statistically significant with all classes of micronutrients including vitamins, minerals, amino acids, and trace elements.

Quantitative analysis by Pearson’s correlation showed serum concentration of cholesterol had a strong positive correlation with serum levels of Vit E (r=0.4086, p<0.0001), Vit D3 (r=0.5125, p<0.001), and Vit K1 (r=0.2334, p<0.0001) The cholesterol levels are also considerably correlated with Vit A and iron (<0.05). The serum levels of cholesterol were found to have a significantly negative correlation with folate and asparagine. LDL was found to have a significant positive correlation with serum levels of Vit E (r=0.3398, p<0.0001) and Vit D3 (r=0.4774, p<0.0001).

Pearson’s correlation was carried out to analyze the univariant relationship between serum lipids, lipoproteins, and micronutrients with significance set at *p*<0.05. Significant results are shown in Table 3. The directionality of the variation of micronutrients is represented in Fig 2. It was interesting to note the significant association of branched-chain amino acids with several markers in the cardiovascular panel. Increased levels of BCAA (branched-chain amino acids) Leucine, Isoleucine, and Valine showed significant association with decreased levels of HDL and increased levels of triglycerides Fig 1. Increased levels of Valine were shown to have a significant association with increased levels of LDL and Apo B. These associations show the significant association between branched-chain amino acids and lipid dysregulation. Additionally, amino acids asparagine and glutamine were found to be inversely correlated with serum LDL. A significant inverse correlation was observed between glutamine and serine with serum levels of Apo B.

**Figure 1.**
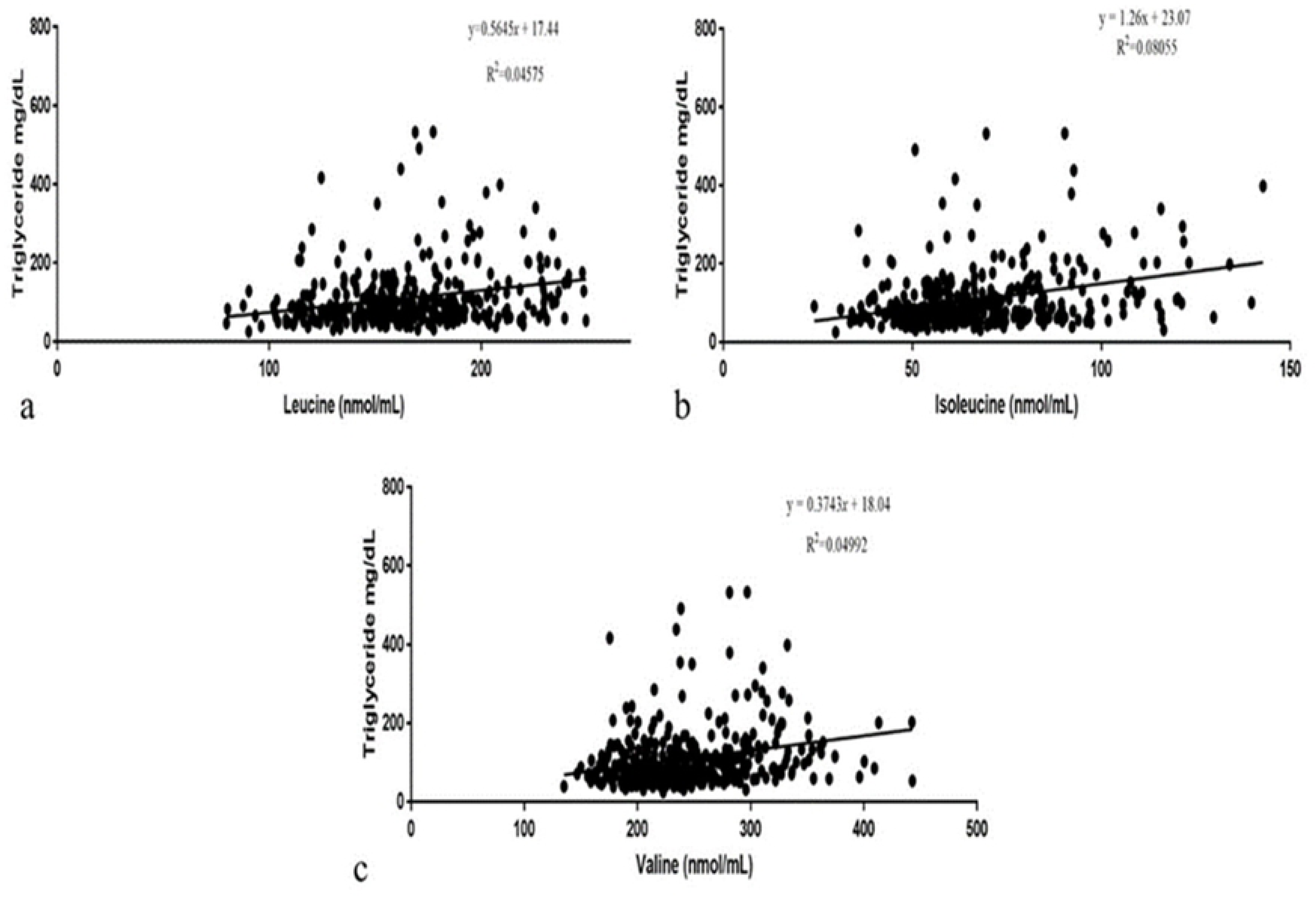
Correlation between serum triglycerides with serum BCAA a. Leucine, b. Isoleucine, c. Valine

**Table 3:**
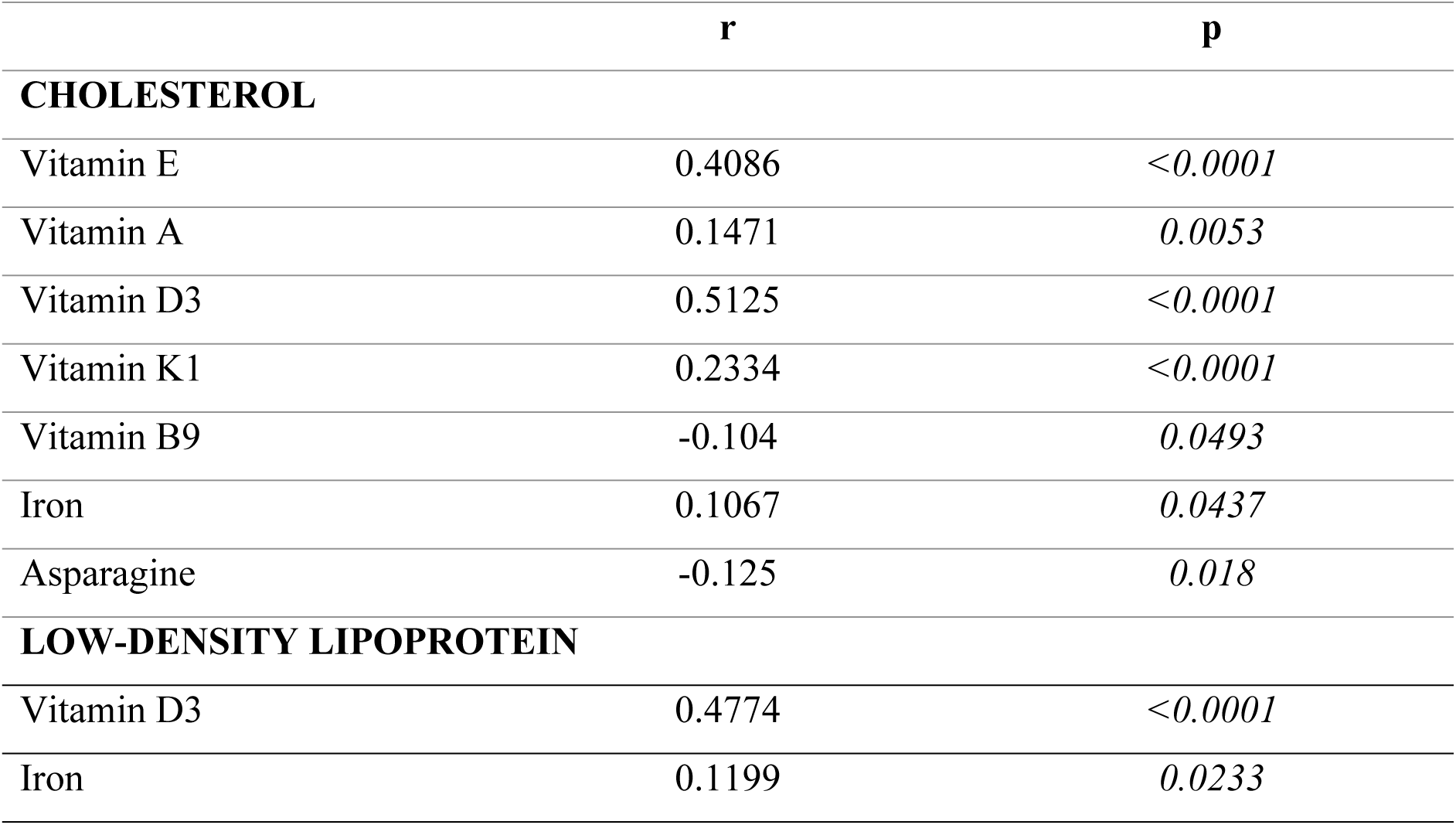
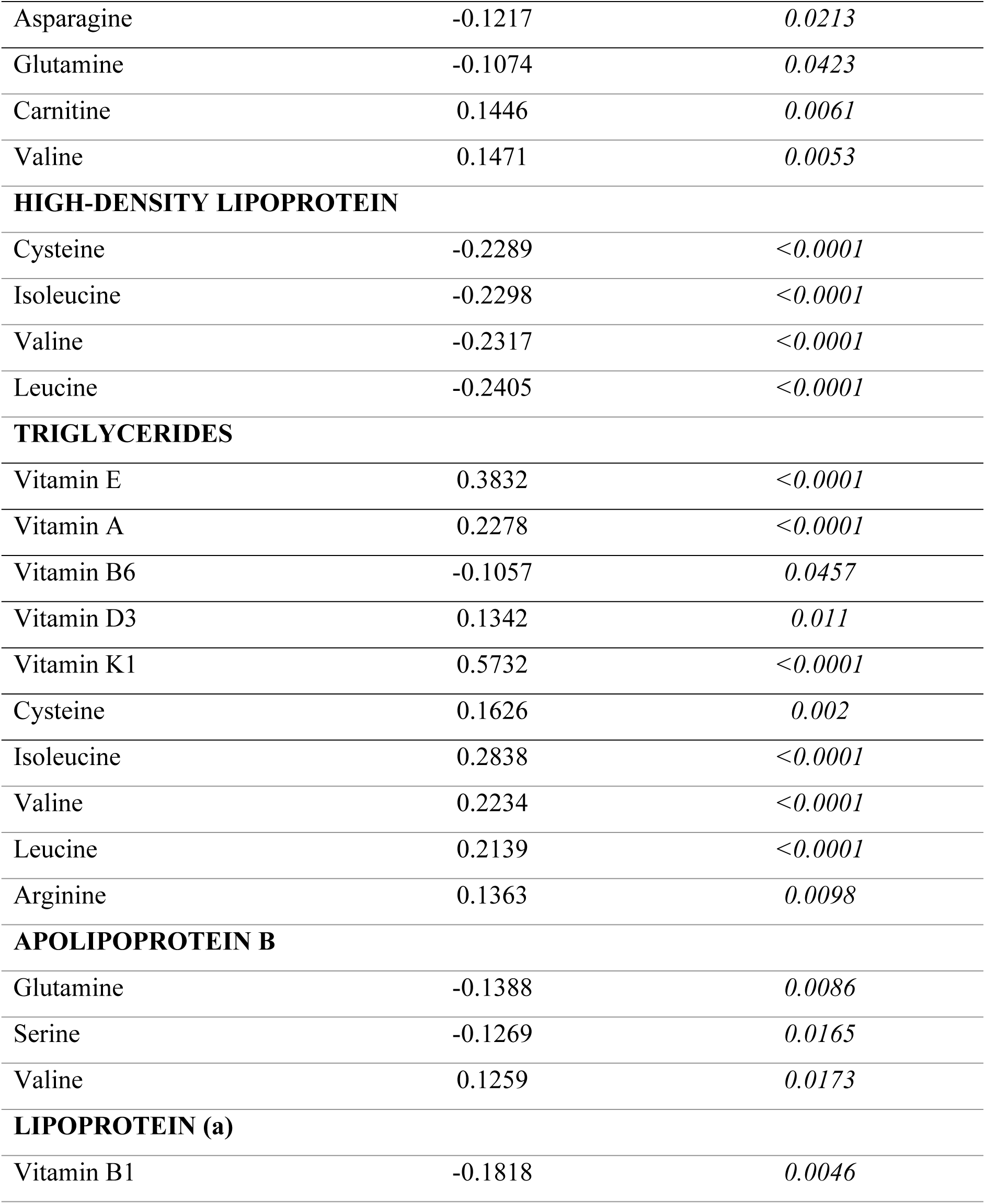
Pearson correlation between micronutrients with serum lipids and lipoproteins

## Discussion

Micronutrient deficiency is a major cause of the pathogenesis, progression, mortality, and morbidity in developing various chronic health issues including CVD. It is regarded as a vital component of the socio-economic development of a society and hence effective strategies have been framed to address deficiencies at the population level by adding emphasis on dietary improvement, supplementation, food fortification, and global public health control. Understanding the role of individual micronutrients and proper intake of the essential micronutrients plays a vital role in preventing micronutrient-related diseases. Micronutrient deficiency due to malnutrition can be prevented by the above-mentioned strategies. But in the context of the current study, the knowledge on the role of individual micronutrients and timely dietary supplementation of micronutrients plays a crucial role in improving physiological functions [11]. Micronutrients such as vitamins, minerals, and amino acids are known to improve cardiovascular health by regulating the serum levels of lipids and lipoproteins. However, the association of micronutrients with cardiovascular markers remains unclear [12]. The present study details the vital micronutrients with significant association with primary cardiovascular markers such as cholesterol, LDL, HDL, triglycerides, Apo A, Apo B, and Lp(a). The study specifies a wide range of micronutrients which includes 13 vitamins, 9 minerals, and 8 amino acids.

The results of the present study signify a strong positive correlation with high significance between fat-soluble vitamins Vit A, Vit D3, Vit E, and Vit K1 with cholesterol and triglycerides. Few studies have discussed the association of fat-soluble vitamins with lipid profile, Piran et al. [13] attempted to study the association of fat-soluble vitamins A, E, and D. They reported a correlation only between vitamin E and lipid profile, they also suggested the supplementation of vitamin E for overweight subjects to decrease LDL-C levels. Several substantial shreds of evidence have proved that significant changes in mean serum retinol levels resulting from dietary intake or when supplemented to reduce the risk of cancer alter lipid metabolism by increasing circulating levels of triglycerides and cholesterol. Pastorino et al. [14] have stated retinol-induced liver damage by abnormal elevation of alkaline phosphatase with overconsumption of vitamin A. Several reports have demonstrated the association of vitamin D with serum lipid profiles, a study by Dibada [15] reported an inverse correlation where the supplementation of vitamin D resulted in reducing serum cholesterol, LDL, and triglycerides, but they reported no association between vitamin D and HDL. A randomized trial by Barbarawi et al. [16] observed no association between vitamin D supplementation and reduced risk of major adverse cardiovascular events, myocardial infarction, stroke, etc. In the present result, vitamin D was found to have a positive correlation with cholesterol and triglycerides. As Barbarawi et al. [16] stated the present result also showed no association with serum HDL. At optimal levels of serum concentrations, vitamin D might be beneficial by reducing cholesterol, LDL, and triglycerides. High doses of Vit D supplements result in irregular heartbeat and can also raise blood calcium levels which lead to heart failure (Galior et al., 2018). An increase in Vit E might interfere with blood clotting by inhibiting platelet aggregation and Vitamin K-mediated clotting factors which might lead to hemorrhagic stroke (Natural Medicines Comprehensive Database). Vitamin E below the reference range results in acute heart muscle damage. Vitamins B9 (folate) were found to have a negative correlation with cholesterol and Vit D3 was found to have a negative correlation with serum LDL. Interestingly no vitamins exhibited any significant correlation with HDL and Apo A. The current results signify that an overdose of fat-soluble vitamins could result in adverse health effects by modulating serological biomarkers. Water-soluble vitamins tend to accumulate less in tissues as they are readily soluble in water and excreted [18]. Among all the analyzed minerals, iron was found to be positively correlated with serum levels of cholesterol (r=0.1067, p<0.0437) and LDL (r=0.1199, p<0.0233).

Numerous studies have demonstrated the significance of micronutrients on cardiovascular health. For instance, Ma et al. [19] suggested that the deficiency in levels of trace minerals such as zinc, copper, iron, and selenium are directly associated with cardiovascular diseases, and supplementation of these trace elements can prevent CVD. A review by Panchal et al. [20] has demonstrated the importance of selenium, vanadium, and chromium in improving metabolic syndrome. Li et al. [21] in their study on plasma metabolomics stated that plasma metabolites and serum micronutrient levels can be used to predict the risk of CVD. Extensive research has increased the understanding of the association between micronutrients and cardiac health. Micronutrients such as vitamins and minerals are widely investigated, whereas amino acids are hardly investigated. A recent report by Tharrey et al. [22] has detailed the association between amino acid intake and cardiovascular mortality and they also reported the negative effect of a few amino acids on cardiac health. The intake of protein and peptide-based health supplements has increased in recent years which could result in increased levels of non-essential amino acids, the effect of this on lipid and lipoprotein levels was analyzed in our study.

Levels of a few vital minerals such as magnesium, potassium, calcium, and zinc have also been known to influence cardiac health. For instance, deficiency in levels of magnesium and potassium results in elevated blood pressure. Calcium levels need to be at an optimum since the deficiency causes pulmonary hypertension, while the excess results in plaque build-up in arteries.

To our knowledge, the present study is the first to report on the association of various amino acids with serum lipid and lipoprotein components. The study observed a statistical significance between various amino acids and serum levels of lipids and lipoproteins as represented in Fig 2. Increased levels of asparagine and glutamine were shown to decrease the levels of LDL. Deprivation of asparagine and glutamine results in lethal effects on endothelial cells (EC). The supplementation of these non-essential amino acids can assist in the recovery of EC, can also fight against nitrification stress, and aid in angiogenesis. A previous report by Luo et al. [23] on the interactive effects of asparagine and aspartate stated that the subjects with type 2 diabetes were observed to have low levels of HDL-C and reported a negative correlation between serum levels of asparagine and HDL-C. Serum levels of glutamine and serine were found to have no influence on plasma lipids and also regular dietary supplementation of these amino acids has various clinical advantages [24]. A study by Mansour et al. [25] demonstrated the advantage of glutamine supplementation as it remarkably reduced the total cholesterol and blood pressure in patients with type 2 diabetes. They also proposed that supplementation of glutamine can be an effective pharmaconutrient in controlling diabetes, obesity, and other chronic metabolic disorders. Similarly, adequate supplementation of serine could help in increased antioxidant and anti-fatty streak (lesion in the development of atherosclerosis) activity. The increase in the serum levels of triglycerides is significantly associated with low serum levels of amino acids such as glutamine, serine, leucine, isoleucine, and valine. An increase in the serum levels of triglycerides results in atherosclerosis which leads to high chances of stroke and other related heart diseases [26]. A significant positive association was observed between elevated levels of Apo B and low serum levels of amino acids such as serine, and isoleucine. Serum levels of Apo B above adequate levels are associated with the accumulation of LDL-C and non-HCL-C which are associated with various CVD.

**Figure 2.**
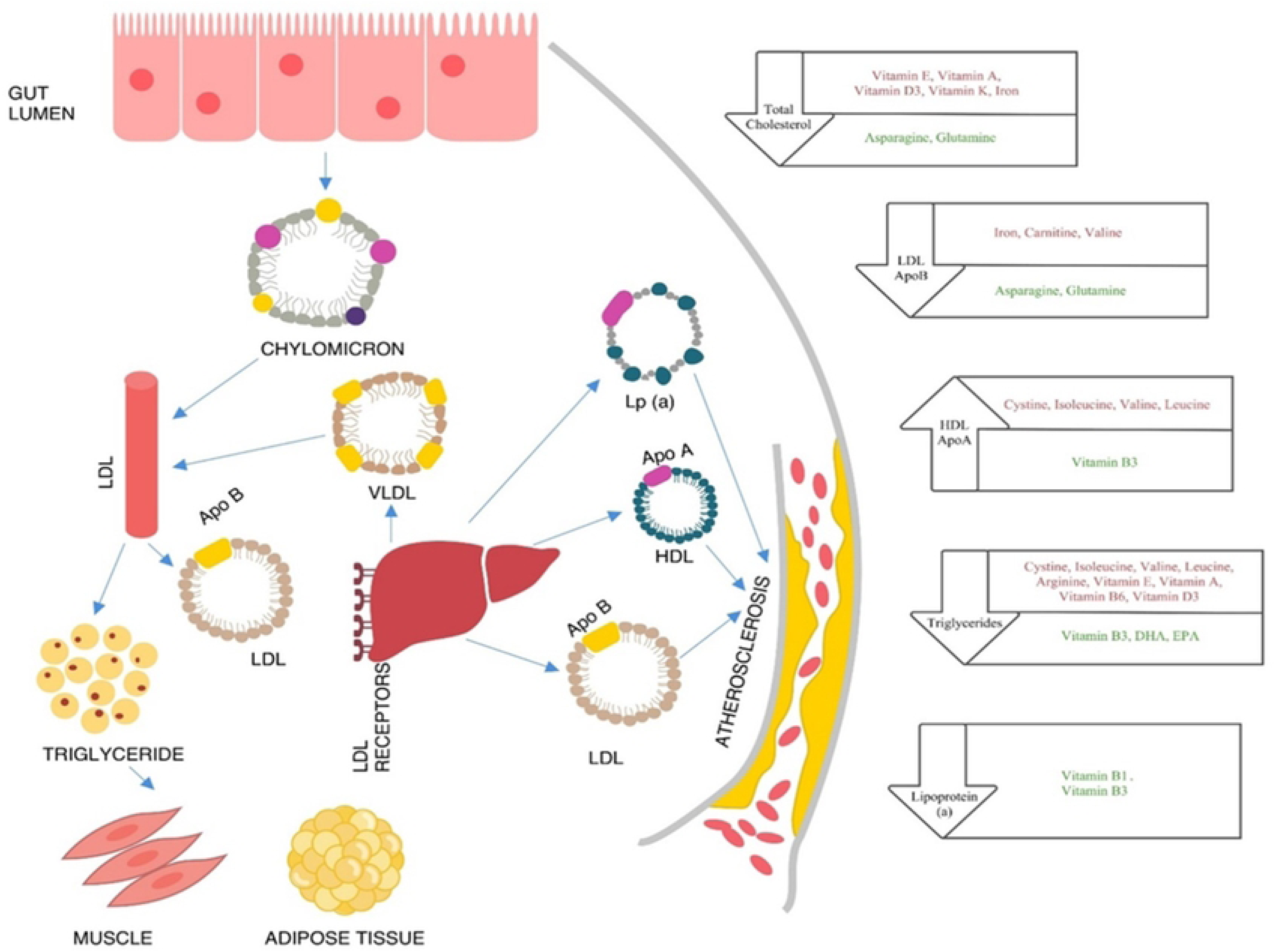
Role of micronutrients in regulating serum lipids Micronutrients in green font reduce the risk by modulating the particular component of the lipid panel, Micronutrients in red font increase the risk by modulating the particular component of the lipid panel

The present study brings out an interesting correlation between cardiovascular markers and BCAA’s (branched-chain amino acids). BCAA’s, leucine, isoleucine, and valine are nonpolar, and hydrophobic, and are vital nitrogen sources for the synthesis of glutamine and alanine [27]. Several epidemiological studies have proved the association between BCAAs and increased risk of type 2 diabetes [28]. The increase in the levels of BCAA’s increases the oxidation of BCAA’s in muscles which inhibits the fatty acid oxidation and results in blunted insulin signaling. Diabetes and CVD are the major causes of morbidity and mortality. A report by Magnusson et al., [28] identified BCAA as a strong predictor of CVD development and also as an early marker for the association between diabetes and CVD. Another study by Tobias et al., [29] proved, that circulating BCAA’s as strong predictors of type 2 diabetes. The quantitative analysis of BCAA’s - leucine, isoleucine, and valine with the cardio markers showed a clear association. A strong negative correlation between HDL and BCAAs and a strong positive correction between triglycerides and BCAAs (Table 3). This signifies the increase in the serum level of BCAA decreases the HDL and increases the triglycerides. While in the case of LDL and Apo B a positive correlation was observed with serum level of valine which signifies the increase in the serum levels of valine also increases the levels of LDL and Apo B which could result in CVD.

The present results identify a few micronutrients with the possibility of being pathogenic, for instance, the positive correlation between vitamin D and serum cholesterol and LDL. The same association has been previously reported by numerous studies including a report by Giri et al. [30]. Grimes [31] has reported that in the absence of sunlight the inactive vitamin D is diverted to the synthesis of cholesterol. Even so, intestinal absorption is the only source of fat-soluble vitamins and their mechanism of intestinal absorption remains unclear. Recent studies revealed the participation of several membrane proteins in cholesterol absorption evidenced by advancements in genome-editing, genome-wide association, and gene mutation analysis on cholesterol and intense studies in cholesterol absorption inhibitors. Interestingly, these analyses also revealed that cholesterol transporters can also transport fat-soluble vitamins [32]. In association with these studies, the present study identifies the serum levels of fat-soluble vitamins as effective markers of elevated cholesterol transportation and can be used as an early predictor of CVD. It’s a well-established fact that increased serum BCAA was associated with elevated triglycerides and reduced HDL [33]. The present results also identify a similar pathology between serum HDL, Triglycerides and BCAA. The BCAA in the present study exhibited a significant negative correlation with HDL and a strong positive correlation with triglycerides. Consumption of energy-dense and high-palatable foods has increased in recent years, and consumption of BCAAs is one such major behavioral change received considerable attention in recent times. The most recent study by Latimer et al. (2021) reported that an increase in BCAA consumption resulted in the dramatic growth of the heart and also increased the progression of cardiac diseases.

One of the main limitations of our study is that it includes data from free-living people with limited information on diet and lifestyle choices. The current study signifies the association of vital cardio markers with a diverse class of micronutrients. Further research is required for a better understanding of the clinical significance of these micronutrients and also to understand the biochemical interference of vitamins, minerals, and amino acids in lipid metabolism.

## Conclusions

The current study examines the explicit correlation of micronutrients with various cardio markers. Given the broad set of micronutrients evaluated their significance in regulating vital cardio markers was better understood. The study highlights the negative correlation of various vitamins and amino acids on cardiovascular health which was a significant observation. The deficiency of various micronutrients leads to a significant increase in cardiovascular disease risk due to dysregulation of lipid and lipoprotein markers. Similarly increased levels of certain micronutrients especially branched-chain amino acids lead to increased cardiovascular risk due to lipid and lipoprotein dysregulation. It is important to look at both deficiencies and overconsumption of micronutrients to optimize nutrient intake. The study suggests the proper monitoring of body vitals and optimal intake of micronutrients are to be considered for their effective roles in metabolic pathways to implement a risk reduction strategy for cardiovascular health.

## Data Availability

To access supporting data, contact the bioinformatics team at or the corresponding author at.

## Acknowledgments

We thank Qingqing Yue, Junior Graphic Designer for assisting with designing figures and Vibrant America LLC for supporting this research.

## Statement of Ethics

An ethics statement was not required for this study type

## Author Contributions

Hari Krishnamurthy, Karthik Krishna, and Tianhao Wang performed the research. Hari Krishnamurthy, John J. Rajasekaran, Karenah Rajasekaran, and Vasanth Jayaraman designed the study. Qi Song, Kang Bei, and Swarnkumar Reddy analyzed the data. Hari Krishnamurthy and Swarnkumar Reddy wrote the article.

## Institutional Review Board Statement

IRB exemption (work order #1-1098539-1) was determined by the Western Institutional Review Board (WIRB) for Vibrant America Biorepository to use de-linked and deidentified remnant human specimen and medical data for research purposes.

## Data Availability

All relevant data are within the manuscript and its Supporting Information files. Any additional data will be available upon request from Vibrant America LLC by sending an email to our bioinformatics team at

## Supplementary Material

**Table S1:** Association of serum cholesterol with micronutrients

|  | Greater than Reference<br>Range (n=62) |  | Within range (n=278) |  | P<br>(P<0.05) |
| --- | --- | --- | --- | --- | --- |
| ASPARAGINE | 53.8±12.7 | 51.4(34.1-93.1) | 54.1±12.7 | 52.9(28-113.0) | 0.8067 |
| GLUTAMINE | 505.7±82.7 | 512.8(210-752) | 504.2±80.6 | 503.1(202.2-741) | 0.8270 |
| SERINE | 144.9±34.3 | 138(75-213.4) | 149.3±35.3 | 146.1(59.6-282.9) | 0.2869 |
| CYSTEINE | 17.3±8.7 | 17.0(2.2-46.2) | 17.7±8.9 | 16.2 (1.3-45.2) | 0.8426 |
| SELENIUM | 139±32 | 140.1(-47-207.5) | 143.3±30.5 | 139 (93.4-360.4) | 0.8771 |
| VITAMIN E | 16±5.7 | 14.4 (7.9-29.4) | 12.9±4.2 | 11.9 (5.3-30.7) | 0.0001 |
| CHOLINE | 14.4±5.6 | 13.3(6.1-35.1) | 14.1±5.7 | 12.5(1.0-39.6) | 0.5605 |
| CARNITINE | 28.8±7.7 | 29.1 (12.6-44.3) | 26.5±7.1 | 26.6(4.1-50.5) | 0.0317 |
| SODIUM | 141.9±2.9 | 142 (137-148) | 140.1±22.0 | 141 (-222-155) | 0.2091 |
| POTASSIUM | 4.5±0.4 | 4.6 (3.1-5.6) | 4.5±0.47 | 4.4 (3.72-6.22) | 0.1658 |
| CALCIUM | 9.8±0.3 | 9.8(9-10.6) | 9.6±0.4 | 9.6 (8.5-11.2) | 0.0001 |
| MANGANESE | 0.6±0.2 | 0.6 (0.3-1.7) | 0.7±0.4 | 0.6 (0.2-6.8) | 0.6281 |
| ZINC | 0.7±0.1 | 0.7 (0.4-.9) | 0.7±0.1 | 0.6 (0.3-2.2) | 0.0160 |
| COPPER | 1.0±0.2 | 0.9 (0.6-2.1) | 1.05±0.3 | 1.0 (0.3-3.4) | 0.7309 |
| CHROMIUM | 0.2±0.1 | 0.2 (0.05-0.6) | 0.2±0.1 | 0.22 (0.01-1.7) | 0.6393 |
| IRON | 108.7±33.3 | 102.0 (34.8-206.5) | 101.9±38.8 | 99.9 (25.3-238.9) | 0.1217 |
| MAGNESIUM | 2.2±0.1 | 2.2 (1.9-2.6) | 2.1±0.1 | 2.1 (1.0-2.6) | 0.0001 |
| VITAMIN A | 80.7±25.1 | 77.8(34.1-142.9) | 76.0±23.6 | 71.5 (37.4-151.9) | 0.0942 |
| VITAMIN_B1 | 21.3±13.5 | 19.4 (1.0-70.3) | 21.6±15.0 | 18.3 (0.8-136.8) | 0.8489 |
| VITAMIN_B2 | 26.8±25.3 | 19.4 (6.2-174.1) | 28.1±21.9 | 21.9 (4.8-140.8) | 0.4462 |
| VITAMIN_B3 | 17.4±7.2 | 18.1 (3.4-34.6) | 19.7±8.8 | 19.1 (2.7-110.1) | 0.0559 |
| VITAMIN_B6 | 15.3±13.0 | 11.3 (1.1-70.8) | 17.5±19.6 | 12.3 (1.6-236.7) | 0.4704 |
| VITAMIN_B12 | 787.9±358.1 | 758 (245.1-1958) | 719±319 | 650 (196-1923) | 0.1395 |
| VITAMIN_B5 | 99.6±74.4 | 70.8 (23.5-315.2) | 107.5±89.2 | 71.0 (13.8-482.1) | 0.6705 |
| VITAMIN_C | 0.4±0.2 | 0.4 (.05-1.2) | 0.4±0.3 | 0.3 (.05-3.74) | 0.5634 |
| VITAMIN_D3 | 1.1±0.2 | 1.1 (0.4-1.7) | 0.8±0.2 | 0.8 (0.3-1.6) | 0.0001 |
| VITAMIN_K1 | 2.1±1.5 | 1.5 (0.1-7.3) | 1.4±1.3 | 1.2 (0.05-13.9) | 0.0003 |
| VITAMIN_K2 | 0.8±1.7 | 0.2 (0.11-11.2) | 0.6±1.0 | 0.2 (0.1-11.0) | 0.1909 |
| FOLATE | 13.6±4.4 | 13.6(2.9-19.9) | 13.6±3.9 | 13.9 (2.8-19.9) | 0.8426 |
| VITAMIN D 25 OH | 50.2±26.6 | 41.2 (11.4-133) | 46.5±22.5 | 40.9 (8.8-136) | 0.5307 |
| ISOLEUCINE | 68.0±17.2 | 66.9 (35.7-115.7) | 68.4±21.1 | 64.6 (24.0-142.7) | 0.6996 |
| VALINE | 255.7±54.7 | 248.3 (146.5-400) | 242.3±55.4 | 233.2 (134.9-442) | 0.0484 |
| LEUCINE | 166.5±37.4 | 159.0 (79.7-247.6) | 162.6±34.2 | 159.1 (80.3-249.2) | 0.5826 |
| CITRULLINE | 31.4±7.6 | 31.3 (17.3-52.1) | 29.6±8.1 | 28.8 (10.1-69.3) | 0.0774 |
| ARGININE | 121.8±32 | 117.5 (65.1-220.6) | 123.2±33.8 | 119.1 (49.6-299.2) | 0.7459 |

**Table S2:** Association of serum LDL with micronutrients

|  | Greater than Reference<br>Range (n=146) |  | Within range (n=212) |  | P<br>(P<0.05) |
| --- | --- | --- | --- | --- | --- |
| ASPARAGINE | 52.9±11.2 | 51.7(31.1-108.0) | 54.8±12.7 | 53.1(28.1-113.0) | 0.2238 |
| GLUTAMINE | 496.9±80.5 | 502.3(204.1-752.4) | 510.6±80.3 | 507.6(202.2-741.1) | 0.1594 |
| SERINE | 148.4±33.9 | 144.1(75-227.9) | 148.4±35.4 | 146.4(59.6-282.9) | 0.9280 |
| CYSTEINE | 18.5±8.7 | 17.9(2.8-46.2) | 17.3±9.1 | 15.6(1.3-45.2) | 0.1121 |
| SELENIUM | 142.2±23.6 | 139.4(93.4-210.8) | 143.6±32.1 | 141.4(95.7-360.4) | 0.8131 |
| VITAMIN E | 14.8±4.8 | 13.6(7.3-29.4) | 12.3±4.2 | 11.2(5.3-30.7) | 0.0001 |
| CHOLINE | 14.2±5.6 | 13.0(5.8-39.6) | 14.0±5.7 | 12.6(1.0-36.6) | 0.7838 |
| CARNITINE | 28.0±7.3 | 28.3(10.2-50.5) | 26.1±7.1 | 26.3(4.1-43.6) | 0.0258 |
| SODIUM | 141.7±3.4 | 142 (133-151) | 139.7±25.1 | 141(129-155) | 0.3443 |
| POTASSIUM | 4.5±0.3 | 4.5(3.1-5.6) | 4.5±0.4 | 4.5(3.6-6.2) | 0.7340 |
| CALCIUM | 9.6±0.3 | 9.6 (8.5-10.6) | 89.6±0.4 | 9.6(8.6-11.2) | 0.5248 |
| MANGANESE | 0.69±0.26 | 0.63(0.2-1.85) | 0.74±0.54 | 0.63(0.3-6.82) | 0.9838 |
| ZINC | 0.72±0.12 | 0.71(0.39-1.39) | 0.70±0.18 | 0.67(0.44-2.23) | 0.0373 |
| COPPER | 1.05±0.29 | 1.02(0.37-2.53) | 1.05±0.31 | 1.0 (0.4-3.4) | 0.8362 |
| CHROMIUM | 0.2±0.1 | 0.2(0.02-1.0) | 0.26±0.17 | 0.2(0.011-1.79) | 0.4714 |
| IRON | 108±33.3 | 108.1(35.5-201.8) | 99.1±41.4 | 91.4(25.3-238.9) | 0.0024 |
| MAGNESIUM | 2.23±0.16 | 2.2(1.5-2.66) | 2.16±0.19 | 2.1(1.0-2.6) | 0.0002 |
| VITAMIN A | 78.3±22.7 | 74.6(34.1-151.5) | 76.2±24.8 | 72.2(37.1-151.9) | 0.2012 |
| VITAMIN_B1 | 20.8±13.5 | 18.0(1.0-75.9) | 22.4±15.9 | 18.8(0.83-136.8) | 0.4553 |
| VITAMIN_B2 | 27.2±22.8 | 21.1(5.7-174.1) | 29.2±25.8 | 21.6(4.8-222.5) | 0.7505 |
| VITAMIN_B3 | 19.4±10.5 | 18.6(3.4-110.1) | 19.0±6.7 | 18.2(2.7-40.6) | 0.7766 |
| VITAMIN_B6 | 16.0±13.2 | 12.2(1.1-84.2) | 18.2±21.3 | 12.3(1.4-236.7) | 0.6179 |
| VITAMIN_B12 | 744.3±328.8 | 657.2(245.1-1958) | 726.1±323.4 | 663.9(196-1923) | 0.6856 |
| VITAMIN_B5 | 103.6±87.5 | 70.6(13.8-482.1) | 111.1±9.2 | 71.5(19.2-407.4) | 0.5429 |
| VITAMIN_C | 0.45±0.29 | 0.41(0.05-2.62) | 0.45±0.31 | 0.39(0.08-3.74) | 0.7418 |
| VITAMIN_D3 | 1.04±0.26 | 1.02(0.44-1.78) | 0.84±0.81 | 0.81(0.32-1.63) | 0.0001 |
| VITAMIN_K1 | 1.65±1.2 | 1.29(0.05-7.32) | 1.56±1.4 | 1.25(0.15-13.9) | 0.3875 |
| VITAMIN_K2 | 0.68±1.2 | 0.28(0.10-11.2) | 0.72±1.17 | 0.29(0.082-11.0) | 0.5063 |
| FOLATE | 13.8±4.0 | 14.1(2.9-19.9) | 13.6±3.9 | 13.9(2.8-19.9) | 0.5133 |
| VITAMIN D 25 OH | 47.8±22.6 | 41.5(10.4-133) | 46.3±23.5 | 40.5(8.8-136) | 0.3757 |
| ISOLEUCINE | 69.0±20.1 | 67.0(31.0-139.7) | 68.6±21.0 | 64.4(24.0-142.7) | 0.6712 |
| VALINE | 250.5±55.1 | 242.0(146.5-442.7) | 241.1±54.2 | 232.5(134.9-442.4) | 0.0903 |
| LEUCINE | 166.4±35.1 | 159.3(79.7-248.2) | 161.5±34.4 | 158.2(87.7-249.2) | 0.2160 |
| CITRULLINE | 30.1±7.6 | 29.4(10.3-51.8) | 29.9±8.3 | 28.8(10.1-69.3) | 0.7509 |
| ARGININE | 122.2±33.2 | 118.9(61.0-220.6) | 123.3±33.8 | 118.4(49.6-299.2) | 0.7600 |

**Table S3:** Association of serum HDL with micronutrients

|  | Less than Reference<br>Range (n=36) |  | Within range (n=301) |  | P<br>(P<0.05) |
| --- | --- | --- | --- | --- | --- |
| ASPARAGINE | 105.6±42.6 | 110.1(34.1-220.6) | 53.9±11.6 | 52.9(28.1-108.0) | 0.0001 |
| GLUTAMINE | 212.5±164.3 | 124.6(61.0-671.4) | 508.6±80.7 | 510.9(202.2-752.4) | 0.0001 |
| SERINE | 128.0±36.2 | 124.7(61.0-282.9) | 149.4±34.2 | 145.6(59.6-274.5) | 0.0001 |
| CYSTEINE | 20.4±7.8 | 20.1(6.7-42.7) | 17.8±9.5 | 16.5(2.5-46.2) | 0.0364 |
| SELENIUM | 141.1±24.8 | 142.2(96.8-210.8) | 143.0±31.5 | 140.6(93.4-360.4) | 0.7682 |
| VITAMIN E | 15.8±5.3 | 16.2(6-30.7) | 13.2±4.5 | 12.1(5.3-29.4) | 0.0011 |
| CHOLINE | 13.3±4.8 | 11.7(6.1-27.3) | 14.2±5.8 | 12.9(1.0-39.6) | 0.3783 |
| CARNITINE | 27.3±6.6 | 26.9(12.8-40.3) | 26.9±7.3 | 27.2(4.1-50.5) | 0.7748 |
| SODIUM | 141.5±3.5 | 142(134-150) | 141.4±3.2 | 141(129-155) | 0.9044 |
| POTASSIUM | 4.4±0.43 | 4.3(3.1-5.3) | 4.5±0.39 | 4.5(3.6-6.2) | 0.2055 |
| CALCIUM | 9.6±0.5 | 9.5(8.8-11.2) | 9.6±0.3 | 9.6(8-11.1) | 0.3304 |
| MANGANESE | 0.70±0.2 | 0.68(0.32-1.2) | 0.72±0.4 | 0.62(0.2-6.8) | 0.3071 |
| ZINC | 0.68±0.1 | 0.6(0.4-0.9) | 0.7±0.1 | 0.6(0.3-2.2) | 0.2098 |
| COPPER | 1.1±0.3 | 1.0(0.4-2.5) | 1.04±0.30 | 0.9(0.3-3.4) | 0.3136 |
| CHROMIUM | 0.23±0.10 | 0.21(0.06-0.52) | 0.2±0.16 | 0.2(0.01-1.7) | 0.6318 |
| IRON | 92.2±33.6 | 86.5(25.3-192.1) | 104.1±38.2 | 100(29.8-238.9) | 0.0983 |
| MAGNESIUM | 2.1±0.2 | 2.1(1.2-2.5) | 2.1±0.18 | 2.2(1.0-2.6) | 0.8566 |
| VITAMIN A | 84.8±27.0 | 80.0(43.3-151.9) | 76.7±23.9 | 72.3(34.1-151.5) | 0.0661 |
| VITAMIN_B1 | 21.1±14.0 | 17.7(1.0-56.8) | 21.4±15.0 | 18.4(0.8-136.8) | 0.8646 |
| VITAMIN_B2 | 21.8±14.3 | 17.2(6.4-64.2) | 28.1±21.8 | 22.0(4.8-140.8) | 0.0848 |
| VITAMIN_B3 | 19.4±5.5 | 19.4(5.4-30.3) | 19.4±8.8 | 18.9(2.7-110.1) | 0.7625 |
| VITAMIN_B6 | 13.5±13.6 | 9.1(2.9-80.9) | 17.6±19.0 | 12.4(1.1-236.7) | 0.1048 |
| VITAMIN_B12 | 642±304.0 | 532.2(245.1-1593) | 742.3±331.1 | 674.3(150-1958) | 0.0509 |
| VITAMIN_B5 | 104.3±84.9 | 71.8(25.8-330.0) | 106.0±87.9 | 68.3(13.8-482.1) | 0.8070 |
| VITAMIN_C | 0.43±0.23 | 0.39(0.08-0.88) | 0.44±0.29 | 0.39(0.05-3.74) | 0.9910 |
| VITAMIN_D3 | 0.93±0.24 | 0.93(0.41-1.46) | 0.92±0.27 | 0.88(0.32-1.78) | 0.5533 |
| VITAMIN_K1 | 2.3±2.7 | 1.6(0.2-13.9) | 1.5±1.0 | 1.2(0.05-7.3) | 0.0290 |
| VITAMIN_K2 | 0.74±0.87 | 0.32(0.10-3.8) | 0.71±1.2 | 0.28(0.082-11.2) | 0.2940 |
| FOLATE | 12.0±4.1 | 11.0(2.9-19.6) | 13.7±3.9 | 13.9(2.8-19.9) | 0.0146 |
| VITAMIN D 25 OH | 40.7±24.9 | 33.6(12.4-136) | 48.3±22.8 | 41.7(8.8-133) | 0.0091 |
| ISOLEUCINE | 76.8±24.3 | 73.2(35.7-142.7) | 67.9±20.2 | 65.1(24.0-139.7) | 0.0193 |
| VALINE | 256.6±57.0 | 247.8(175.3-363.9) | 244.1±54.8 | 234.8(134.9-442.1) | 0.2253 |
| LEUCINE | 169.3±35.8 | 168.7(113.7-241.0) | 163.4±34 | 158.3(80.3-249.2) | 0.3703 |
| CITRULLINE | 28.3±8.3 | 28.7(10.3-44.9) | 30.2±8.0 | 29.1(10.1-69.3) | 0.3014 |
| ARGININE | 129.6±33.0 | 119.5(78.3-205.3) | 122.5±34.1 | 119.2(49.6-299.2) | 0.3083 |

**Table S4:** Association of serum triglycerides with micronutrients

|  | Greater than Reference Range<br>(n=36) |  | Within range (n=323) |  | P<br>(P<0.05) |
| --- | --- | --- | --- | --- | --- |
| ASPARAGINE | 53.4±15.5 | 49.9(31.1-118.7) | 54.3±12.2 | 53.1(28.1-113.0) | 0.3363 |
| GLUTAMINE | 463.7±107.6 | 472.9(118.7-608.3) | 508.4±78.7 | 507.6(202.2-752) | 0.0290 |
| SERINE | 136.5±32.8 | 130.6(76.0-198.5) | 149.6±34.7 | 145.6(59.6-282.9) | 0.0368 |
| CYSTEINE | 22.1±18.1 | 19.4(4.9-118.7) | 17.6±9.1 | 15.7(1.3-46.2) | 0.0573 |
| SELENIUM | 142.6±22.8 | 141(96.8-207.5) | 143.0±29.5 | 140.2(93.4-360.4) | 0.7712 |
| VITAMIN E | 20.1±17 | 16.7(8.5-118.7) | 12.9±4.3 | 11.9(5.3-29.4) | 0.0001 |
| CHOLINE | 17.8±18.1 | 14.1(7.0-118.7) | 14.0±5.7 | 12.6(1.0-39.6) | 0.1661 |
| CARNITINE | 32.6±16.5 | 31.5(11.6-118.7) | 26.5±7.2 | 26.6(4.1-50.5) | 0.0014 |
| SODIUM | 141.5±4.8 | 142.5(118.7-150) | 141.5±3.4 | 141(129-155) | 0.3449 |
| POTASSIUM | 7.7±19.0 | 4.5(4.1-118.7) | 4.5±0.3 | 4.5 (3.1-6.2) | 0.6792 |
| CALCIUM | 12.8±18.1 | 9.7(9.0-118.7) | 9.6±0.4 | 9.6(8.2-11.1) | 0.0518 |
| MANGANESE | 0.7±0.2 | 0.67(0.32-1.5) | 0.72±0.46 | 0.63(0.2-6.8) | 0.1321 |
| ZINC | 0.68±0.13 | 0.702(0.4-1.0) | 0.71±0.16 | 0.69(0.39-2.23) | 0.5508 |
| COPPER | 1.01±0.23 | 1.0(0.48-1.5) | 1.0±0.3 | 1.0(0.3-3.4) | 0.6959 |
| CHROMIUM | 0.23±0.11 | 0.22(0.05-0.51) | 0.25±0.16 | 0.22(0.01-1.7) | 0.6972 |
| IRON | 101.2±29.1 | 97.5(50.7-160) | 102.8±39.5 | 100(25.3-238.9) | 0.9361 |
| MAGNESIUM | 2.2±0.15 | 2.2(1.9-2.6) | 2.1±0.19 | 2.2(1.0-2.6) | 0.7556 |
| VITAMIN A | 88.4±27.3 | 82(52.8-151.9) | 75.8±23.3 | 72.1(34.1-151.5) | 0.0061 |
| VITAMIN_B1 | 19.8±15.7 | 16.9(1.0-66.4) | 21.9±14.9 | 18.6(0.8-136.8) | 0.2701 |
| VITAMIN_B2 | 26.8±15.5 | 24.5(6.2-64.7) | 27.9±22.9 | 20.8(4.8-174.1) | 0.4594 |
| VITAMIN_B3 | 17.5±5.6 | 16.9(8.7-27.8) | 19.4±8.7 | 18.9(2.7-110.1) | 0.1979 |
| VITAMIN_B6 | 13.2±10.8 | 9.8(2.9-49.1) | 17.7±19.0 | 12.4(1.1-236.7) | 0.1156 |
| VITAMIN_B12 | 688.9±321.3 | 552.1(281.5-1593) | 738.2±325.9 | 667.8(196-1958) | 0.2503 |
| VITAMIN_B5 | 112.6±97.3 | 70.5(27.4-437.4) | 107.5±88.3 | 71.2(13.8-482.1) | 0.6561 |
| VITAMIN_C | 0.51±0.42 | 0.41(0.14-2.6) | 0.44±0.28 | 0.40(0.05-3.74) | 0.6053 |
| VITAMIN_D3 | 0.96±0.29 | 0.93(0.40-1.42) | 0.91±0.26 | 0.88(0.32-1.78) | 0.3611 |
| VITAMIN_K1 | 3.14±2.7 | 2.3(0.22-13.9) | 1.42±0.99 | 1.22(0.05-7.32) | 0.0001 |
| VITAMIN_K2 | 1.07±1.9 | 0.32(0.10-11.2) | 0.66±1.09 | 0.28(0.082-11.0) | 0.1134 |
| FOLATE | 12.4±4.5 | 12.6(2.8-19.6) | 13.8±3.9 | 14.1(2.9-19.9) | 0.0839 |
| VITAMIN D 25 OH | 33.9±13.6 | 30.7(15.4-71.2) | 48.6±23.2 | 41.8(8.8-136) | 0.0001 |
| ISOLEUCINE | 84.0±27.5 | 84.2(35.7-142.7) | 67.1±19.1 | 64.6(24.0-139.7) | 0.0002 |
| VALINE | 275.0±63.3 | 281.1(175.3-442.4) | 241.7±52.8 | 233.6(134.9-442.1) | 0.0020 |
| LEUCINE | 179.9±36.8 | 182.7(113.7-233.5) | 161.7±34.1 | 157.4(79.7-249.2) | 0.0033 |
| CITRULLINE | 29.3±7.5 | 30.8(10.3-41.7) | 30.0±8.1 | 28.8(10.1-69.3) | 0.8478 |
| ARGININE | 134.4±38.1 | 125.0(76.9-211.3) | 121.6±32.8 | 117.7(49.6-299.2) | 0.0539 |

**Table S5:**
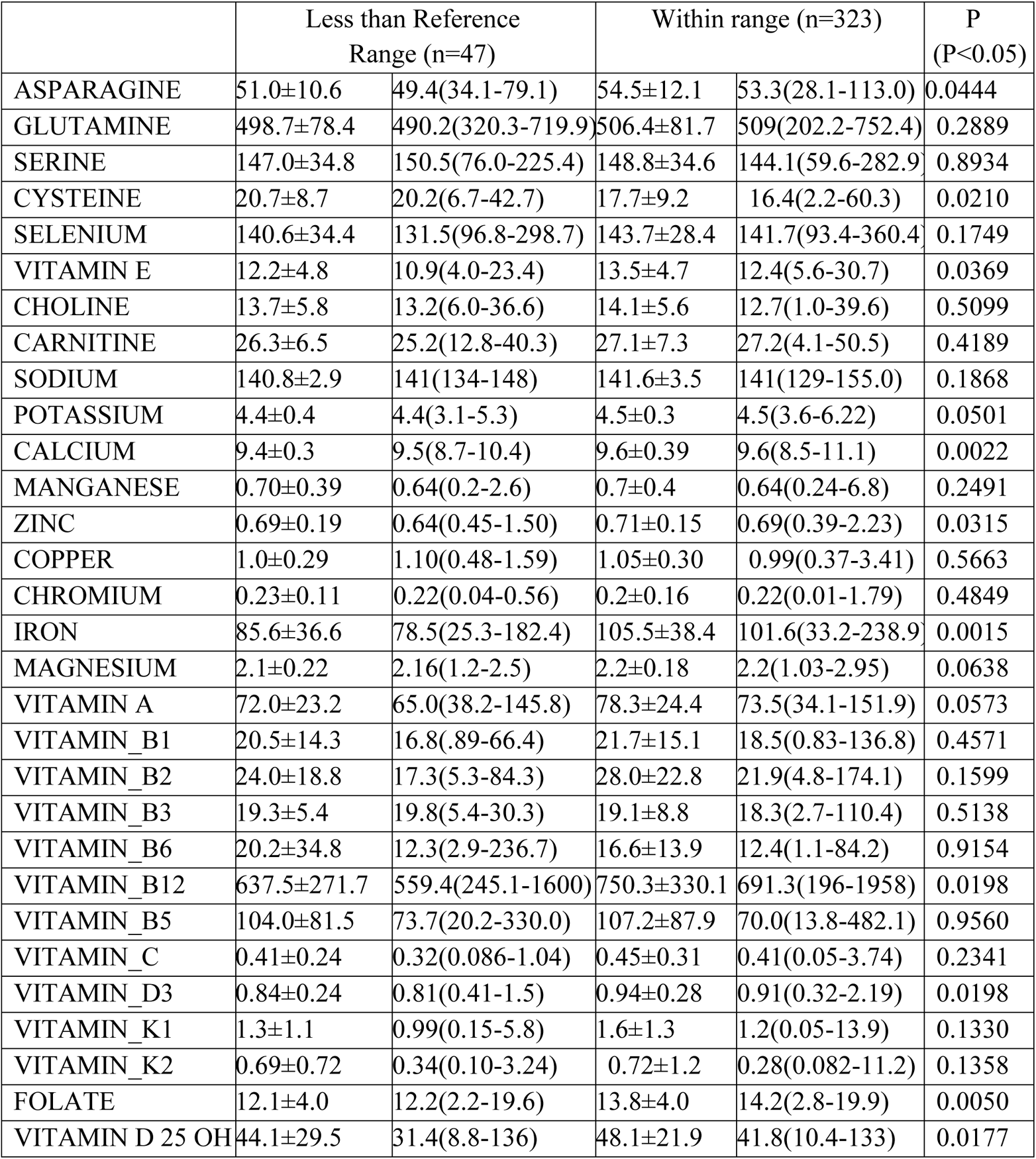
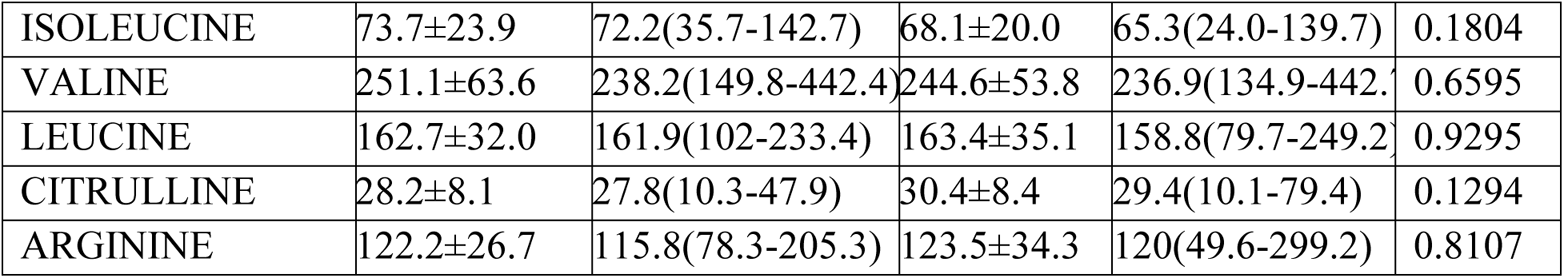
Association of serum Apo A with micronutrients

**Table S6:** Association of serum Apo B with micronutrients

|  | Greater than Reference<br>Range (n=87) |  | Within range (n=270) |  | P<br>(P<0.05) |
| --- | --- | --- | --- | --- | --- |
| ASPARAGINE | 53.4±11.1 | 53.5(34.1-79.1) | 54.2±12.3 | 52.8(28.1-113.0) | 0.7013 |
| GLUTAMINE | 489.6±84.4 | 492.2(204.1-752.4) | 510.5±79.7 | 507.8(202.2-741.0) | 0.0643 |
| SERINE | 140.6±33.0 | 135.4(75.0-209.7) | 151.1±34.7 | 147.2(59.6-282.9) | 0.0189 |
| CYSTEINE | 18.4±9.5 | 17.9(2.8-60.3) | 18.0±9.1 | 16.5(2.2-45.2) | 0.6201 |
| SELENIUM | 146.4±24.0 | 144.8(93.4-210.8) | 142.3±30.3 | 139(95.7-360.4) | 0.0451 |
| VITAMIN E | 16.0±5.5 | 15.3(7.3-29.0) | 12.5±4.1 | 11.7(4.0-30.7) | 0.0001 |
| CHOLINE | 14.4±5.3 | 13.2(6.6-39.6) | 13.9±5.7 | 12.4(1.0-36.6) | 0.3090 |
| CARNITINE | 27.8±7.7 | 28.2(9.8-44.3) | 26.7±7.1 | 26.6(4.1-50.5) | 0.1555 |
| SODIUM | 141.5±3.4 | 141(133-151) | 141.5±3.4 | 141(129-155) | 0.8920 |
| POTASSIUM | 4.5±0.35 | 4.5(3.6-5.6) | 4.5±0.40 | 4.49(3.1-6.22) | 0.0377 |
| CALCIUM | 9.7±0.3 | 9.7(8.7-10.6) | 9.6±0.3 | 9.6(8.5-11.1) | 0.0055 |
| MANGANESE | 0.73±0.29 | 0.67(0.34-2.1) | 0.73±0.49 | 0.63(0.24-6.82) | 0.1635 |
| ZINC | 0.73±0.11 | 0.72(0.39-1.0) | 0.70±0.17 | 0.68(0.44-2.23) | 0.0058 |
| COPPER | 1.0±0.32 | 1.0(0.6-2.5) | 1.0±0.3 | 1.0(0.3-3.4) | 0.4660 |
| CHROMIUM | 0.24±0.11 | 0.22(0.05-0.59) | 0.25±0.17 | 0.22(0.011-1.7) | 0.9988 |
| IRON | 109.3±34.2 | 107.7(45.6-201.3) | 100.8±39.9 | 97.7(25.3-238.9) | 0.0239 |
| MAGNESIUM | 2.2±0.1 | 2.2(1.9-2.9) | 2.1±0.19 | 2.1(1.0-2.6) | 0.0002 |
| VITAMIN A | 82.4±23.7 | 77.7(34.1-145.8) | 75.8±24.2 | 72.1(37.1-151.9) | 0.0096 |
| VITAMIN_B1 | 19.3±14.1 | 16.8(1.17-70.3) | 22.2±15.2 | 18.6(0.83-136.8) | 0.0516 |
| VITAMIN_B2 | 27.1±23.8 | 20.9(6.2-174.1) | 28.5±24.8 | 21.2(4.8-222.5) | 0.7427 |
| VITAMIN_B3 | 18.7±11.9 | 17.8(3.4-110.1) | 19.3±7.0 | 19.1(2.7-40.6) | 0.0954 |
| VITAMIN_B6 | 15.5±14.3 | 11.7(2.9-84.2) | 17.6±19.1 | 12.5(1.1-236.7) | 0.2511 |
| VITAMIN_B12 | 750.2±345.3 | 659.5(290.2-1958) | 730.6±318.6 | 666.8(196-1923) | 0.8363 |
| VITAMIN_B5 | 110.1±89.9 | 72.0(13.8-437.4) | 96.5±76.8 | 68.6(23.5-482.1) | 0.4485 |
| VITAMIN_C | 0.45±0.29 | 0.40(0.05-3.74) | 0.45±0.32 | 0.39(0.05-2.6) | 0.7818 |
| VITAMIN_D3 | 0.87±0.24 | 0.83(0.32-1.6) | 1.09±0.30 | 1.08(0.44-2.19) | 0.0001 |
| VITAMIN_K1 | 1.5±1.3 | 1.2(0.05-13.9) | 1.8±1.4 | 1.4(0.10-7.3) | 0.0416 |
| VITAMIN_K2 | 0.6±1.0 | 0.29(0.082-11.0) | 0.86±1.6 | 0.28(0.10-11.2) | 0.9964 |
| FOLATE | 13.0±4.3 | 13.3(2.8-19.8) | 13.8±3.9 | 14.2(2.2-19.9) | 0.1956 |
| VITAMIN D 25 OH | 47.6±22.3 | 41.7(8.8-136) | 47.5±25.3 | 40.1(15.4-133) | 0.4508 |
| ISOLEUCINE | 72.7±19.9 | 70.7(34.0-121.4) | 67.6±20.7 | 64.2(24.0-142.7) | 0.0138 |
| VALINE | 261.9±55.2 | 262.7(146.5-404.0) | 240.2±54.2 | 232.1(134.9) | 0.0004 |
| LEUCINE | 172±34.8 | 168.8(113.7-248.2) | 160.5±34.2 | 157.4(79.7-249.2) | 0.0138 |
| CITRULLINE | 30.1±8.0 | 28.8(10.1-69.3) | 30.2±9.6 | 29.7(10.3-79.4) | 0.9509 |
| ARGININE | 123.7±33.8 | 119.4(49.6-299.2) | 122.2±32.0 | 117.5(61.1-211.3) | 0.7427 |

**Table S7:** Association of serum Lipoprotein A with micronutrients

|  | Greater than Reference<br>Range (n=87) |  | Within range (n=270) |  | P<br>(P<0.05) |
| --- | --- | --- | --- | --- | --- |
| ASPARAGINE | 54.0±10.0 | 54.2(31.1-80.6) | 53.5±12.3 | 52.7(28.1-108.0) | 0.4975 |
| GLUTAMINE | 508.3±81.7 | 514.8(320.3-710.3) | 505.3±80.5 | 506.9(202.2-741.0) | 0.8216 |
| SERINE | 151.4±35.3 | 150.7(59.6-274.5) | 148.0±37.8 | 142.5(70.5-241.1) | 0.4179 |
| CYSTEINE | 17.9±8.6 | 7.02(2.8-45.1) | 19.0±9.9 | 17.4(3.3-45.2) | 0.6336 |
| SELENIUM | 139.2±21.9 | 138.3(96.6-210.8) | 141.5±25.9 | 138.0(93.4-274.7) | 0.7976 |
| VITAMIN E | 13.5±4.8 | 12.0(5.6-29.4) | 13.1±4.2 | 12.4(5.3-28.2) | 0.9456 |
| CHOLINE | 13.6±5.6 | 11.9(5.8-36.6) | 14.6±6.1 | 13.2(1.0-39.6) | 0.1288 |
| CARNITINE | 26.5±7.1 | 26.7(12.5-50.5) | 26.8±7.7 | 27.0(4.1-44.3) | 0.7048 |
| SODIUM | 141.5±3.5 | 142(129-150) | 141.6±3.6 | 141(133-155) | 0.7881 |
| POTASSIUM | 4.5±0.4 | 4.5(3.1-5.6) | 4.5±0.39 | 4.4(3.7-6.2) | 0.4339 |
| CALCIUM | 9.6±0.4 | 9.5(8.7-10.6) | 9.6±0.4 | 9.6(8.5-10.8) | 0.6777 |
| MANGANESE | 0.70±0.29 | 0.64(0.24-1.85) | 0.74±0.32 | 0.69(0.3-2.6) | 0.2023 |
| ZINC | 0.72±0.15 | 0.69(0.45-1.5) | 0.7±0.14 | 0.69(0.3-1.3) | 0.4235 |
| COPPER | 1.0±0.29 | 1.0(0.10-2.1) | 1.0±0.29 | 1.0(0.5-2.5) | 0.4000 |
| CHROMIUM | 0.24±0.12 | 0.23(0.02-0.68) | 0.25±0.14 | 0.22(0.02-1.0) | 0.8911 |
| IRON | 100.8±38.4 | 95(33.7-238.9) | 105.7±38.4 | 102.0(25.3-201.3) | 0.3270 |
| MAGNESIUM | 2.1±0.9 | 2.1(1.2-2.6) | 2.1±0.2 | 2.1(1.0-2.6) | 0.4372 |
| VITAMIN A | 76.8±25.4 | 71.2(34.3-151.2) | 77.9±25.9 | 71.5(36.9-151.5) | 0.7935 |
| VITAMIN_B1 | 16.8±11.1 | 14.9(1.0-49.2) | 22.7±13.7 | 20.7(0.8-78.8) | 0.0008 |
| VITAMIN_B2 | 26.7±20.4 | 20.3(5.7-135.7) | 30.1±30.4 | 22.2(5.7-222.5) | 0.4808 |
| VITAMIN_B3 | 19.9±6.9 | 19.8(3.4-35.4) | 19.7±10.2 | 18.9(2.7-110.1) | 0.3823 |
| VITAMIN_B6 | 17.2±22 | 11.9(1.6-236.7) | 17.1±16.9 | 11.0(1.4-84.2) | 0.6969 |
| VITAMIN_B12 | 684.3±264.5 | 599.1(196-1381) | 776.9±364.0 | 713.1(200-1923) | 0.1376 |
| VITAMIN_B5 | 103.8±81.9 | 68.6(13.8-360.9) | 101.8±86.0 | 70.0(21.5-482.1) | 0.8976 |
| VITAMIN_C | 0.43±0.24 | 0.38(0.08-1.2) | 0.44±0.33 | 0.4(0.05-3.7) | 0.8100 |
| VITAMIN_D3 | 0.93±0.28 | 0.91(0.3-1.7) | 0.94±0.26 | 0.94(0.32-1.6) | 0.4137 |
| VITAMIN_K1 | 1.5±1.1 | 1.2(0.05-5.8) | 1.5±1.1 | 1.32(0.1-7.3) | 0.5896 |
| VITAMIN_K2 | 0.69±1.2 | 0.29(0.08-11.2) | 0.61±0.78 | 0.28(0.10-4.8) | 0.9556 |
| FOLATE | 13.5±4.2 | 13.7(2.9-19.9) | 13.8±3.7 | 13.9(3.4-19.8) | 0.7113 |
| VITAMIN D 25 OH | 46.7±21.1 | 40.9(10.4-128) | 47.7±23.4 | 41.2(12.4-133) | 0.9703 |
| ISOLEUCINE | 68.8±19.7 | 66.3(33.5-129.7) | 68.6±21.6 | 65.6(24.0-142.7) | 0.8235 |
| VALINE | 243.9±56.8 | 236.2(149.8-442.7) | 243.1±53.4 | 240.0(146.5-409.1) | 0.9007 |
| LEUCINE | 164.4±33.4 | 164.5(90.3-235.1) | 161.1±33.9 | 157.2(80.3-248.2) | 0.3296 |
| CITRULLINE | 29.6±7.8 | 28.6(10.1-51.2) | 30.1±9.2 | 29.6(10.3-69.3) | 0.5236 |
| ARGININE | 124.7±39.2 | 118.3(56.1-299.2) | 125.4±32.8 | 122.4(49.6-220.6) | 0.5700 |

**Table S8:** Pearson correlation between serum cholesterol and micronutrients

|  | r | p |
| --- | --- | --- |
| ASPARAGINE | -0.125 | 0.018 |
| GLUTAMINE | -0.1127 | 0.0331 |
| SERINE | -0.02601 | 0.6238 |
| CYSTEINE | -0.01392 | 0.793 |
| SELENIUM | 0.02659 | 0.616 |
| VITAMIN E | 0.4086 | <0.0001 |
| CHOLINE | 0.03297 | 0.5341 |
| CARNITINE | 0.09912 | 0.061 |
| SODIUM | 0.01349 | 0.7992 |
| POTASSIUM | 0.009154 | 0.863 |
| CALCIUM | 0.01211 | 0.8193 |
| MANGANESE | -0.08336 | 0.1154 |
| ZINC | -0.06639 | 0.2101 |
| COPPER | -0.05695 | 0.2826 |
| CHROMIUM | -0.07196 | 0.1743 |
| IRON | 0.1067 | 0.0437 |
| MAGNESIUM | 0.01114 | 0.8337 |
| VITAMIN A | 0.1471 | 0.0053 |
| VITAMIN_B1 | -0.08282 | 0.1178 |
| VITAMIN_B2 | -0.02302 | 0.6643 |
| VITAMIN_B3 | -0.03355 | 0.5268 |
| VITAMIN_B6 | -0.08714 | 0.0997 |
| VITAMIN_B12 | 0.03583 | 0.4992 |
| VITAMIN_B5 | -0.0739 | 0.1629 |
| VITAMIN_C | -0.02218 | 0.6757 |
| VITAMIN_D3 | 0.5125 | <0.0001 |
| VITAMIN_K1 | 0.2334 | <0.0001 |
| VITAMIN_K2 | 0.07045 | 0.1835 |
| FOLATE | -0.104 | 0.0493 |
| VITAMIN_D_25_OH | 0.06205 | 0.2416 |
| ISOLEUCINE | 0.007779 | 0.8834 |
| VALINE | 0.09774 | 0.0647 |
| LEUCINE | 0.04598 | 0.3857 |
| CITRULLINE | 0.02438 | 0.6458 |
| ARGININE | 0.02322 | 0.6614 |

**Table S9:**
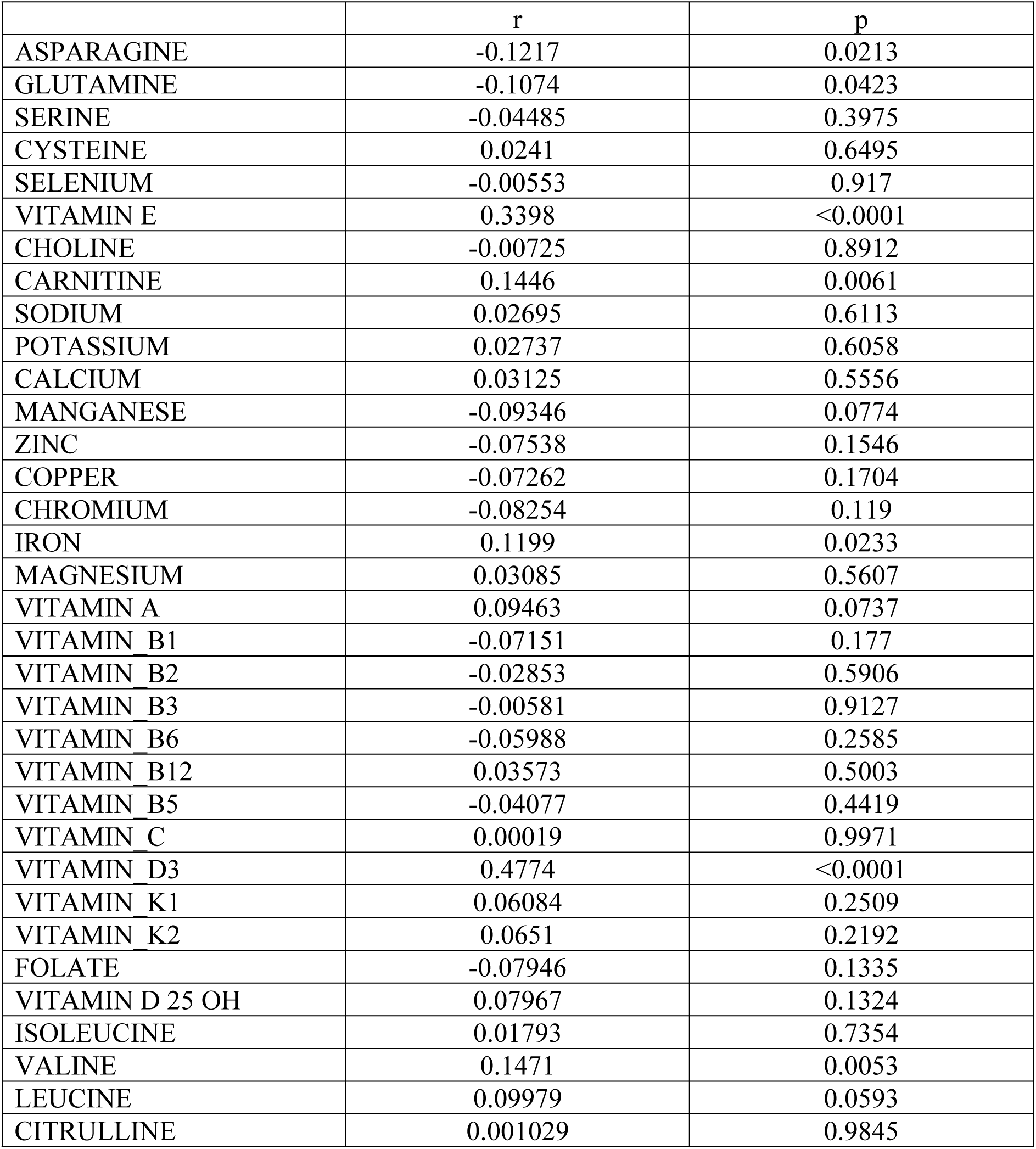
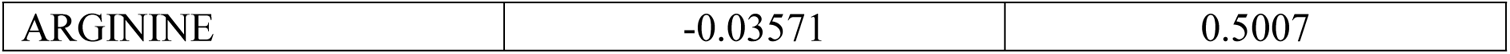
Pearson correlation between serum LDL and micronutrients

**Table S10:** Pearson correlation between serum HDL and micronutrients

|  | r | p |
| --- | --- | --- |
| ASPARAGINE | 0.007988 | 0.8803 |
| GLUTAMINE | 0.01765 | 0.7392 |
| SERINE | 0.06766 | 0.2016 |
| CYSTEINE | -0.2289 | <0.0001 |
| SELENIUM | 0.08967 | 0.0902 |
| VITAMIN E | -0.099 | 0.0613 |
| CHOLINE | 0.01779 | 0.7373 |
| CARNITINE | -0.1653 | 0.0017 |
| SODIUM | -0.08206 | 0.1212 |
| POTASSIUM | -0.0799 | 0.1313 |
| CALCIUM | -0.08061 | 0.1279 |
| MANGANESE | 0.03657 | 0.4903 |
| ZINC | 0.03521 | 0.5067 |
| COPPER | 0.04878 | 0.3574 |
| CHROMIUM | 0.0416 | 0.4326 |
| IRON | 0.03515 | 0.5074 |
| MAGNESIUM | -0.08087 | 0.1267 |
| VITAMIN A | -0.03156 | 0.5517 |
| VITAMIN_B1 | 0.02135 | 0.6873 |
| VITAMIN_B2 | 0.02877 | 0.5874 |
| VITAMIN_B3 | 0.01155 | 0.8276 |
| VITAMIN_B6 | -0.01147 | 0.8288 |
| VITAMIN_B12 | 0.07736 | 0.1441 |
| VITAMIN_B5 | -0.01955 | 0.7123 |
| VITAMIN_C | -0.01446 | 0.7852 |
| VITAMIN_D3 | 0.02496 | 0.6378 |
| VITAMIN_K1 | -0.04944 | 0.3509 |
| VITAMIN_K2 | -0.0078 | 0.8831 |
| FOLATE | -0.02846 | 0.5915 |
| VITAMIN_D_25_OH | 0.0655 | 0.2163 |
| ISOLEUCINE | -0.2298 | <0.0001 |
| VALINE | -0.2317 | <0.0001 |
| LEUCINE | -0.2405 | <0.0001 |
| CITRULLINE | 0.0138 | 0.7948 |
| ARGININE | -0.03223 | 0.5432 |

**Table S11:**
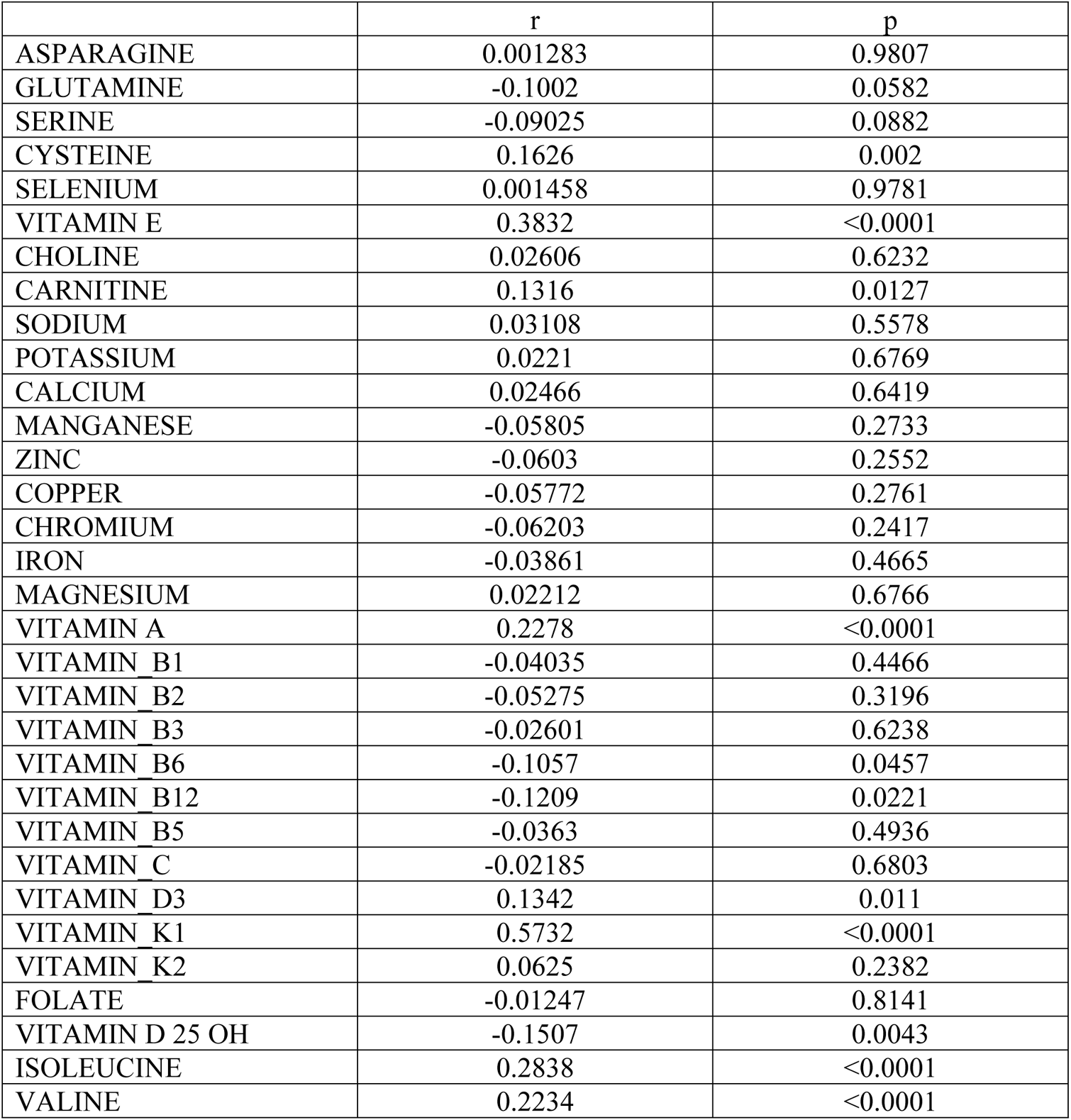
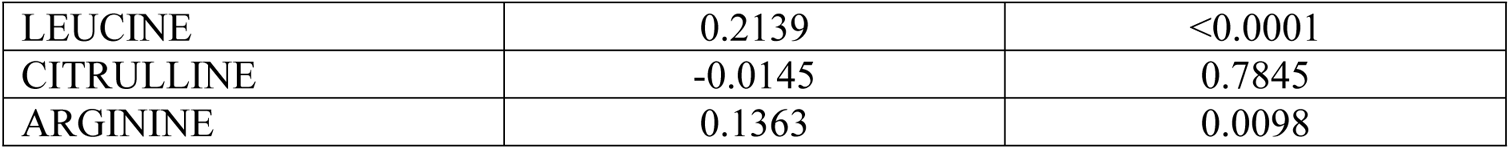
Pearson correlation between serum triglycerides and micronutrients

|  | r | p |
| --- | --- | --- |
| ASPARAGINE | 0.001283 | 0.9807 |
| GLUTAMINE | -0.1002 | 0.0582 |
| SERINE | -0.09025 | 0.0882 |
| CYSTEINE | 0.1626 | 0.002 |
| SELENIUM | 0.001458 | 0.9781 |
| VITAMIN E | 0.3832 | <0.0001 |
| CHOLINE | 0.02606 | 0.6232 |
| CARNITINE | 0.1316 | 0.0127 |
| SODIUM | 0.03108 | 0.5578 |
| POTASSIUM | 0.0221 | 0.6769 |
| CALCIUM | 0.02466 | 0.6419 |
| MANGANESE | -0.05805 | 0.2733 |
| ZINC | -0.0603 | 0.2552 |
| COPPER | -0.05772 | 0.2761 |
| CHROMIUM | -0.06203 | 0.2417 |
| IRON | -0.03861 | 0.4665 |
| MAGNESIUM | 0.02212 | 0.6766 |
| VITAMIN A | 0.2278 | <0.0001 |
| VITAMIN_B1 | -0.04035 | 0.4466 |
| VITAMIN_B2 | -0.05275 | 0.3196 |
| VITAMIN_B3 | -0.02601 | 0.6238 |
| VITAMIN_B6 | -0.1057 | 0.0457 |
| VITAMIN_B12 | -0.1209 | 0.0221 |
| VITAMIN_B5 | -0.0363 | 0.4936 |
| VITAMIN_C | -0.02185 | 0.6803 |
| VITAMIN_D3 | 0.1342 | 0.011 |
| VITAMIN_K1 | 0.5732 | <0.0001 |
| VITAMIN_K2 | 0.0625 | 0.2382 |
| FOLATE | -0.01247 | 0.8141 |
| VITAMIN_D_25_OH | -0.1507 | 0.0043 |
| ISOLEUCINE | 0.2838 | <0.0001 |
| VALINE | 0.2234 | <0.0001 |
| LEUCINE | 0.2139 | <0.0001 |
| CITRULLINE | -0.0145 | 0.7845 |
| ARGININE | 0.1363 | 0.0098 |

**Table S12:** Pearson correlation between serum Apo A and micronutrients

|  | r | p |
| --- | --- | --- |
| ASPARAGINE | -0.04291 | 0.419 |
| GLUTAMINE | 0.003761 | 0.9435 |
| SERINE | 0.0321 | 0.5455 |
| CYSTEINE | -0.05867 | 0.2689 |
| SELENIUM | 0.0395 | 0.4569 |
| VITAMIN E | -0.05202 | 0.327 |
| CHOLINE | -0.03578 | 0.5004 |
| CARNITINE | -0.0026 | 0.9609 |
| SODIUM | -0.06147 | 0.2467 |
| POTASSIUM | -0.0526 | 0.3216 |
| CALCIUM | -0.05579 | 0.2931 |
| MANGANESE | 0.05431 | 0.3062 |
| ZINC | 0.04085 | 0.4417 |
| COPPER | 0.05533 | 0.2971 |
| CHROMIUM | 0.05078 | 0.3387 |
| IRON | 0.01219 | 0.8185 |
| MAGNESIUM | -0.05607 | 0.2907 |
| VITAMIN A | 0.03416 | 0.52 |
| VITAMIN_B1 | 0.009489 | 0.8582 |
| VITAMIN_B2 | -0.05013 | 0.345 |
| VITAMIN_B3 | -0.04226 | 0.426 |
| VITAMIN_B6 | -0.05052 | 0.3412 |
| VITAMIN_B12 | 0.03913 | 0.4611 |
| VITAMIN_B5 | -0.05052 | 0.3412 |
| VITAMIN_C | -0.00363 | 0.9455 |
| VITAMIN_D3 | -0.05824 | 0.2724 |
| VITAMIN_K1 | -0.04603 | 0.3859 |
| VITAMIN_K2 | -0.04088 | 0.4413 |
| FOLATE | 0.07275 | 0.1702 |
| VITAMIN_D_25_OH | -0.07913 | 0.1356 |
| ISOLEUCINE | -0.04946 | 0.3514 |
| VALINE | -0.02571 | 0.6283 |
| LEUCINE | -0.06696 | 0.2069 |
| CITRULLINE | 0.002819 | 0.9577 |
| ARGININE | 0.003076 | 0.9538 |

**Table S13:** Pearson correlation between serum Apo B and micronutrients

|  | r | p |
| --- | --- | --- |
| ASPARAGINE | -0.03841 | 0.4694 |
| GLUTAMINE | -0.1388 | 0.0086 |
| SERINE | -0.1269 | 0.0165 |
| CYSTEINE | -0.02353 | 0.6577 |
| SELENIUM | -0.01502 | 0.7774 |
| VITAMIN E | 0.06915 | 0.1924 |
| CHOLINE | 0.032 | 0.5467 |
| CARNITINE | 0.03729 | 0.4825 |
| SODIUM | 0.04303 | 0.4176 |
| POTASSIUM | 0.03938 | 0.4582 |
| CALCIUM | 0.03969 | 0.4547 |
| MANGANESE | -0.00535 | 0.9198 |
| ZINC | -0.00317 | 0.9525 |
| COPPER | 0.01326 | 0.8028 |
| CHROMIUM | -0.00639 | 0.9042 |
| IRON | 0.08265 | 0.119 |
| MAGNESIUM | 0.04038 | 0.4469 |
| VITAMIN A | 0.1008 | 0.057 |
| VITAMIN_B1 | -0.02215 | 0.6767 |
| VITAMIN_B2 | -0.0354 | 0.505 |
| VITAMIN_B3 | -0.09294 | 0.0795 |
| VITAMIN_B6 | -0.02192 | 0.6798 |
| VITAMIN_B12 | 0.003217 | 0.9517 |
| VITAMIN_B5 | -0.00175 | 0.9737 |
| VITAMIN_C | 0.01495 | 0.7784 |
| VITAMIN_D3 | 0.04051 | 0.4455 |
| VITAMIN_K1 | -0.00555 | 0.9167 |
| VITAMIN_K2 | -0.07411 | 0.1623 |
| FOLATE | 0.1001 | 0.0589 |
| VITAMIN D 25 OH | -0.01943 | 0.7144 |
| ISOLEUCINE | 0.09702 | 0.0671 |
| VALINE | 0.1259 | 0.0173 |
| LEUCINE | 0.03799 | 0.4743 |
| CITRULLINE | -0.06951 | 0.1901 |
| ARGININE | -0.1126 | 0.0334 |

**Table S14:** Pearson correlation between serum lipoprotein A and micronutrients

|  | r | p |
| --- | --- | --- |
| ASPARAGINE | -0.01713 | 0.7914 |
| GLUTAMINE | 0.06125 | 0.3437 |
| SERINE | -0.07783 | 0.2287 |
| CYSTEINE | 0.03708 | 0.5668 |
| SELENIUM | 0.04334 | 0.5031 |
| VITAMIN E | 0.008081 | 0.9007 |
| CHOLINE | -0.06643 | 0.3044 |
| CARNITINE | -0.05322 | 0.4108 |
| SODIUM | -0.03759 | 0.5614 |
| POTASSIUM | 0.09305 | 0.1498 |
| CALCIUM | 0.000307 | 0.9962 |
| MANGANESE | -0.01672 | 0.7963 |
| ZINC | 0.03884 | 0.5485 |
| COPPER | 0.04358 | 0.5007 |
| CHROMIUM | -0.01997 | 0.7577 |
| IRON | 0.07018 | 0.2778 |
| MAGNESIUM | 0.01067 | 0.8691 |
| VITAMIN A | 0.0261 | 0.6869 |
| VITAMIN_B1 | -0.1818 | 0.0046 |
| VITAMIN_B2 | 0.02598 | 0.6882 |
| VITAMIN_B3 | -0.01185 | 0.8548 |
| VITAMIN_B6 | -0.00366 | 0.955 |
| VITAMIN_B12 | -0.08534 | 0.1867 |
| VITAMIN_B5 | 0.01869 | 0.7729 |
| VITAMIN_C | 0.08509 | 0.188 |
| VITAMIN_D3 | 0.05586 | 0.388 |
| VITAMIN_K1 | 0.004489 | 0.9447 |
| VITAMIN_K2 | -0.04037 | 0.5328 |
| FOLATE | -0.03829 | 0.5541 |
| VITAMIN D 25 OH | 0.07921 | 0.2205 |
| ISOLEUCINE | -0.02975 | 0.6458 |
| VALINE | -0.0444 | 0.4927 |
| LEUCINE | 0.02835 | 0.6614 |
| CITRULLINE | -0.02425 | 0.708 |
| ARGININE | -0.0665 | 0.3039 |

